# Childhood interstitial lung disease – application of a stepwise diagnostic classification

**DOI:** 10.1101/2025.09.14.25335703

**Authors:** Christina K Rapp, Matthias Griese

## Abstract

Childhood interstitial lung disease (chILD) represents a heterogeneous group of rare pulmonary disorders. Practical diagnostic approaches tested for feasibility and impact in comprehensive cohorts are lacking.

Objective of this study was to assess a simple etiologically focused classification approach, clarify the role of genetic testing, and quantify the impact of non-pulmonary organ manifestations. We hypothesized that chILD can be classified in a clinically meaningful and versatile way by answering three questions: Which children have an etiological chILD diagnosis due to (1) identified (exposure-related) cause/lung injury, or (2) systemic disease? (3) In how many children without etiological diagnosis can a genetic cause be identified? We also calculated the predictive value of non-pulmonary organ involvement for underlying systemic conditions.

Among 1,686 patients, 24.9% were grouped as ILD with an identified cause/lung injury, 21.6% as ILD with a convincing diagnosis of systemic disease, 18.1% as ILD with genetic diagnosis of systemic disease, 10.1% as ILD with genetic diagnosis affecting the lungs only, and 25.3% as ILD without genetic diagnosis. The average genetic diagnostic yield was 52.8%, with higher rates in interstitial pneumonia (64.0%) or pulmonary alveolar proteinosis (87.2%). The presence of ≥two non-pulmonary organ manifestations increased the likelihood of an underlying systemic disease by two to three-fold. An etiological diagnostic strategy effectively classifies chILD and guides genetic testing. Exome sequencing should be considered if ≥two non-pulmonary organs are involved or if the initial diagnosis becomes uncertain due to an unusual disease course or signs of a second underlying condition.

**Contributor’s statement:** MG managed the project, provided patient’s data, designed data collection software, and monitored data collection. He contributed to the paper through conceptualization and supervised the process, including data curation, formal analysis, validation, statistical analysis and visualizations. He contributed to the writing (original draft, review, and editing) and secured funding. CKR contributed through data curation, formal analysis, investigation, methodology development, validation, analysis, and visualization of the data, as well as writing the original draft, review, and editing. All collaborators of chILD EU registry were involved in the primary data collection process and contributed to the multidisciplinary team responsible for differential diagnosis.

**Funding:** Of relevance for this study were grants from the Deutsche Forschungsgemeinschaft (DFG, Gr970/9-1 and 9-2) Bonn, Germany (Trial registration number: NCT02852928)

**Competing Interests:** All authors have completed the ICMJE uniform disclosure form and declare the following: no support from any organization for the submitted work; no financial relationships with any organizations that might have an interest in the submitted work within the past three years; and receipt of honoraria from Boehringer-Ingelheim for participation in the InPedILD initiative, which did not influence the content or conclusions of the submitted work.

This article has an online data supplement.

## INTRODUCTION

Childhood interstitial lung diseases (chILD), also referred to as pediatric diffuse parenchymal lung diseases (DPLD), represent a wide and diverse spectrum of chronic pulmonary disorders. The currently established classification system stratifies 34 chILD diagnoses, focusing on children younger than two years of age(1), with subsequent expansions incorporating older children(2, 3) and non-pathology-based diagnoses(4). Recent advances in genetic testing often elucidating the precise causes of chILD and generating novel molecularly defined entities(5) challenge current classifications. Prioritizing disease etiologies over phenotypic manifestations(6, 7) appears desirable, however supporting evidence on the applicability of such systems is lacking(8). Subsequent multicenter studies, predominantly retrospective or descriptive, pointed towards the growing role of genetic testing(7, 9–12). Notably, exome or genome sequencing have broadened genetic interrogation, enabling the identification of novel pathogenic variants in previously untargeted genes, either because the gene had not yet been associated with a disease or were not considered due to ambiguous clinical phenotypes. Nevertheless, the genetic spectrum of chILD remains incompletely mapped, and no large study has proven the applicability of a comprehensive standardized diagnostic classification approach.

Thus, this study employed secondary data analysis to apply a primarily etiological categorization of chILD, testing the hypothesis that the systematic application of three key questions allows a meaningful and versatile classification of a large, multidisciplinary team-defined cohort of chILD. We further evaluated the hypothesis that non-pulmonary organ manifestations serve as predictors for an underlying systemic disease. To systematically test this, we employed standardized human phenotype ontology (HPO) terminology(13), ensuring consistency in phenotype characterization while facilitating its computational integration and analysis. Together we quantify the application of an etiological classification system in a large cohort of chILD patients and draw conclusions for clinical practice.

## METHODS

### Study design and participants

This secondary data analysis was conducted at the Childreńs Hospital, Department of Pediatric Pneumology, University of Munich. Ethical approval by Ethics committee of the University Hospital Munich (22–0133, 23–0962, 1503–11). We followed the Declaration of Helsinki and STROBE(14) reporting recommendations facilitating completeness and transparency.

Parents or legal guardians of children participating in the chILD-EU register (www.childeu.net) provided written informed consent for their diagnostic evaluation, processing, storage and usage of their pseudonymized data. For children treated at the pediatric pneumology department in Munich, anonymized data were used as per patient consent and Ethics committee recommendation. Between January’03 and December’24 all pediatric patients submitted to the chILD-EU register and the Munich center with suspected chILD were investigated by an extensive diagnostic work-up, reflecting a standard procedure that has been routinely applied in pediatric pneumology for many years(15). This included the stepwise and if necessary repetitive testing from less to more invasive methods using clinical history and presentation, high-resolution computer tomography imaging, pulmonary function test, biochemical analyses of blood and broncho-alveolar lavage (BAL), genetic testing, and lung histopathology. In patients with suspected pulmonary hypertension, echocardiography or right heart catheterization excluded a cardiac cause. All test results were reviewed by a multidisciplinary team of pediatric pneumologists, pediatric radiologists, geneticists and pathologists, all specialized in chILD. Patients diagnosed with chILD were defined as those presenting for longer than 6 weeks with respiratory complaints, such as tachypnea or dyspnea at rest or with exercise, crackles, retractions, predominantly dry cough, hypoxemia (≤92% oxygen), and diffuse radiological abnormalities. Children with insufficient data or secondary immunodeficiency were excluded (Fig. 1).

**Fig. 1:**
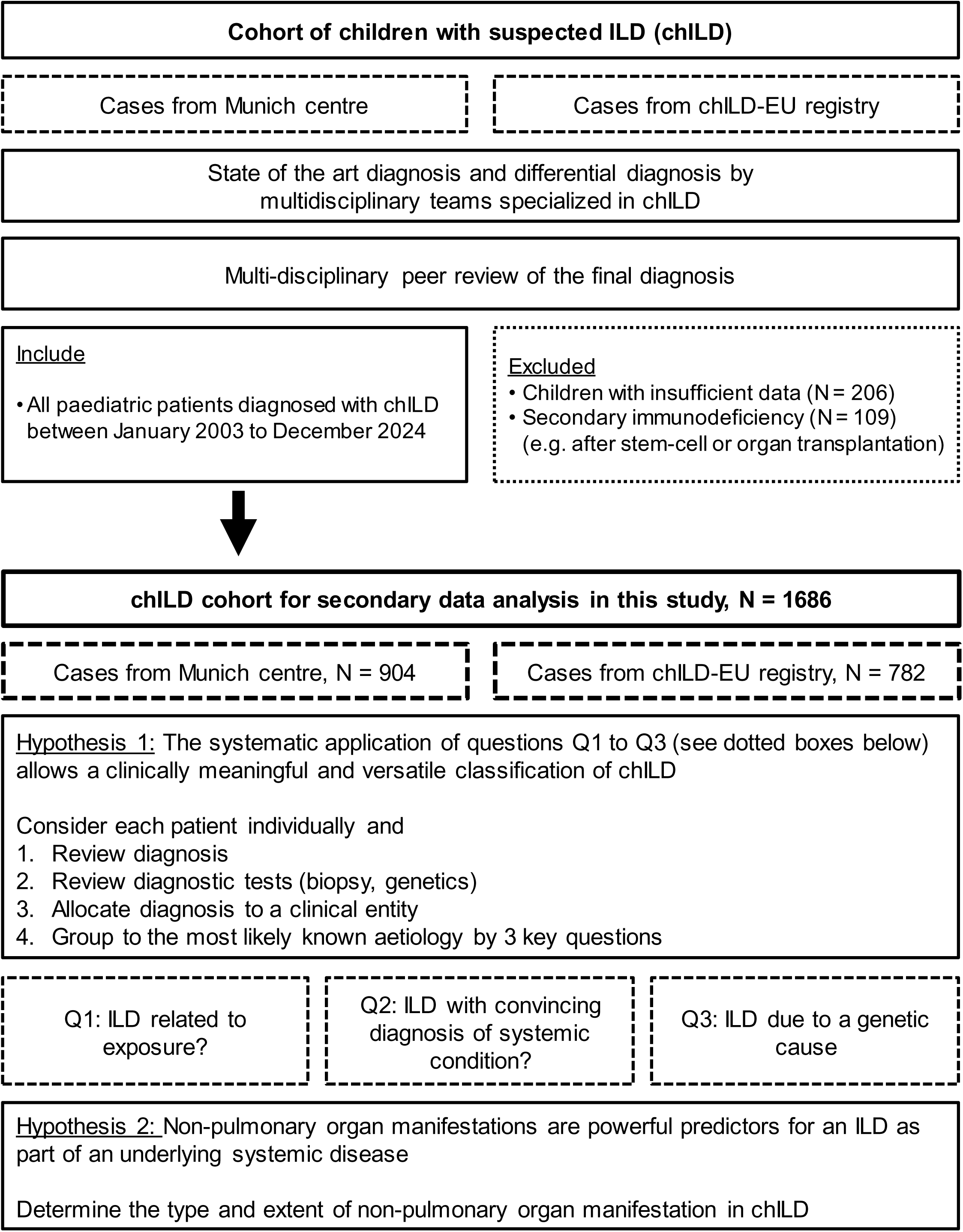
Flow diagram depicting the creation of the chILD cohort investigated, the research hypotheses, and the questions guiding the secondary data analysis.

### Data extraction and secondary data analysis

Data from this cohort, including chILD final differential diagnosis, non-pulmonary organ involvement, genetic data, and histological diagnosis (if performed), were extracted, and utilized for secondary data analysis in the current study(16). Each patient’s data was carefully reviewed and the condition allocated into a pre-genetic clinical entity, utilizing all data, except genetic analysis results.

Then we systematically applied three questions in each patient to achieve an etiology-based classification. First, we asked (question Q1), if we can diagnose an identified (exposure-related) cause/ lung injury responsible for the condition. This included lung prematurity from birth below 30 weeks of gestation and significant respiratory impairment, pulmonary hypoplasia due to lung compression/ displacement, and interstitial pneumonia caused by pathogens like Mycoplasma or non-infection causes due to IgG-mediated hypersensitivity to antigens, inhaled or systemic drug exposures, or chronic aspiration.

Next, each patient was systematically evaluated for systemic disease involvement in the etiology of their ILD. Individuals presenting with a characteristic clinical phenotype consistent with a well-defined condition plausibly accounting for observed ILD manifestations were grouped as having a convincing diagnosis of a systemic disease. These included complex syndromic entities, such as collagen vascular disorders, metabolic and autoimmune conditions, immunodeficiencies, cardiovascular abnormalities, and other multi-organ diseases with established associations to ILD. The term “convincing” reflects the diagnostic likelihood and includes definite diagnoses (probability>90%).

Lastly, the remaining patients and their previously obtained genetic data were evaluated and reviewed for question Q3, if an underlying genetic cause could be identified. The five final etiologic groups are listed with their detailed definition in Tab. S1.

### Genetic testing and variant interpretation

Over time, patients were analyzed by different methods of genetic testing. Initially, genes frequently associated with ILD were analyzed individually, i.e. *ABCA3, SFTPB, and SFTPC* (in this study referred to as Sanger sequencing). Later patients routinely underwent parallel sequencing using next generation sequencing technologies (in this study referred to as panel sequencing). Common genes including *ABCA3, CSF2RA, CSF2RB, FLNA, SFTPB, SFTPC, MARS1, NKX2-1*, and *FOXF1* were analyzed and complemented by *ACVRL1, BMPR2, EIF2AK4, ENG1*, *SMAD1,* and *TBX4,* if pulmonary hypertension was suspected. Since 2019, exome sequencing has been performed more routinely. All genetic variants were interpreted to the current state of knowledge and consistently categorized using ACMG guidelines(17). A molecular genetic diagnosis was considered as identified, if the detected variant was already classified as pathogenic/ likely-pathogenic in a clinical variant database (ClinVar). For novel variants it was essential that the type of consequence, the genotype and observed frequency in control databases (gnomAD) were consistent with linked human disease (OMIM). All identified genetic diagnoses were proven by literature research, if they were previously associated to ILD or other conditions which can cause secondary ILD. Heterozygous variants were regularly confirmed to be absent in healthy parents, except in genes exhibiting incomplete penetrance (e.g., *BMPR2*(*18*), *COPA*(*19*)), where a symptom-free parent could serve as a carrier and low frequency of identified variant (<0.01%) in control databases were tolerated. In cases with two heterozygote variants affecting the same gene, segregation analysis confirmed that these two variants were bi-allelic.

### Evaluation of non-pulmonary organ manifestation

The clinical presentation, particularly non-pulmonary organ involvement, was coded by standardized HPO terms(13). All terms were linked to major organ groups: (1) autoimmune, (2) immunological, (3) hematological, (4) cardiac, (5) vascular, (6) lymphatic, (7) gastrointestinal (including pancreatic), (8) hepatic (including gall bladder), (9) nephrotic (including the urogenital system), (10) skeletal, (11) muscular, (12) neurological, (13) dermal (including hair/ nails), (14) endocrine, and (15) syndromic (including head/ neck) abnormalities. From these, data frequencies of organ involvement and probabilities for the presence of a systemic disease were calculated.

### Statistical analysis

The cohort characteristics were given as median ± IQR or percentage of each cohort. Differences were analyzed by one-way ANOVA (Prism v.9.3.1, significant with p-value <0.05) and post-hoc-tests (Turkey-HSD, SPSS statistics viewer 29.0.0.0). The average number of HPO terms and involved organs were given as mean ± SD, simple and multiple logistic regression were performed (Prism v.9.3.1) and the results were given as odds ratios and presented as 95%CI (profile likelihood). A 95%CI different from 1 was considered statistically significant.

## RESULTS

### Patients’ characteristics and allocation of diagnoses into pre-genetic clinical entities

The database for this study was extracted from a large collection of chILD patients with state of the art’s diagnosis and differential diagnosis conducted by a multidisciplinary team. 1,686 children with confirmed diagnosis of ILD were included. Median age at disease onset was 1.3 years, with a higher prevalence observed in males (Tab. 1, Tab. S2). The study population was characteristic for the entire spectrum of chILD, with 98.2% of the patients originating from Europe. All children underwent a careful individual review of their diagnosis, and the diagnostic tests performed. Their diagnoses were easily and unconstrained allocated into one of 17 pre-genetic clinical entities, reflecting the current disease catalog of the chILD-EU registry. These entities were broadly defined, ordered with increasing complexity of their diagnostic procedures and structured the huge number of different chILD diagnosis in a commonly accepted descriptive manner (Tab. 2).

**Tab. 1.** Characteristics of the study cohort, and patients’ cohorts build in response to question Q1 to Q3, determining the most likely etiology. Data are numbers or median values with first and third quartile. Significant differences between the five etiological groups were calculated by one -way ANOVA and marked by * (p < 0.05) and ** (p < 0.0001)

|  | All patients | ILD with identified cause/ lung injury | ILD with convincing diagnosis of systemic disease | ILD with genetic diagnosis of systemic disease | ILD with genetic diagnosis, affecting lung only | ILD without genetic diagnosis |
| --- | --- | --- | --- | --- | --- | --- |
| Patients, N | 1686 | 419 | 365 | 306 | 170 | 426 |
| ChILD-EU register patients | 782 | 123 | 155 | 180 | 100 | 224 |
| Munich center patients | 904 | 296 | 210 | 126 | 70 | 202 |
| Age at disease onset [years] | 1.3<br>(0.3 to 8.0) | 1.1<br>(0.3 to 8.2) | 5.2**<br>(0.8 to 13.4) | 2.4*<br>(0.5 to 8.0) | 0.4<br>(0.1 to 4.1) | 0.6<br>(0.2 to 2.2) |
| Patients below the age of 2 [%] | 56.1% | 57.8% | 37.3%** | 46.4%* | 67.1%* | 73.2%** |
| Follow-up [years] | 7.1<br>(3.0 to 13.2) | 9.2<br>(5.3 to 15.0) | 7.1<br>(3.7 to 13.5) | 5.2<br>(1.9 to 9.5) | 5.4<br>(0.2 to 8.7) | 7.3<br>(2.5 to 13.3) |
| Male to female ratio | 1 : 0.82 | 1 : 0.62** | 1 : 0.98 | 1 : 0.99 | 1 : 1.05 | 1 : 0.74* |
| Age at death/lung transplantation [years] | 0.5<br>(0.1 to 3.4) | 0.3<br>(0.1 to 0.5) | 2.0*<br>(0.8 to 6.5) | 1.6**<br>(0.3 to 9.7) | 0.2<br>(0.1 to 0.6) | 0.4<br>(0.1 to 3.2) |
| Deceased/lung transplanted [%] | 19.2% | 10.0%** | 12.3% | 25.8% | 41.2%** | 20.4% |

**Tab. 2:**
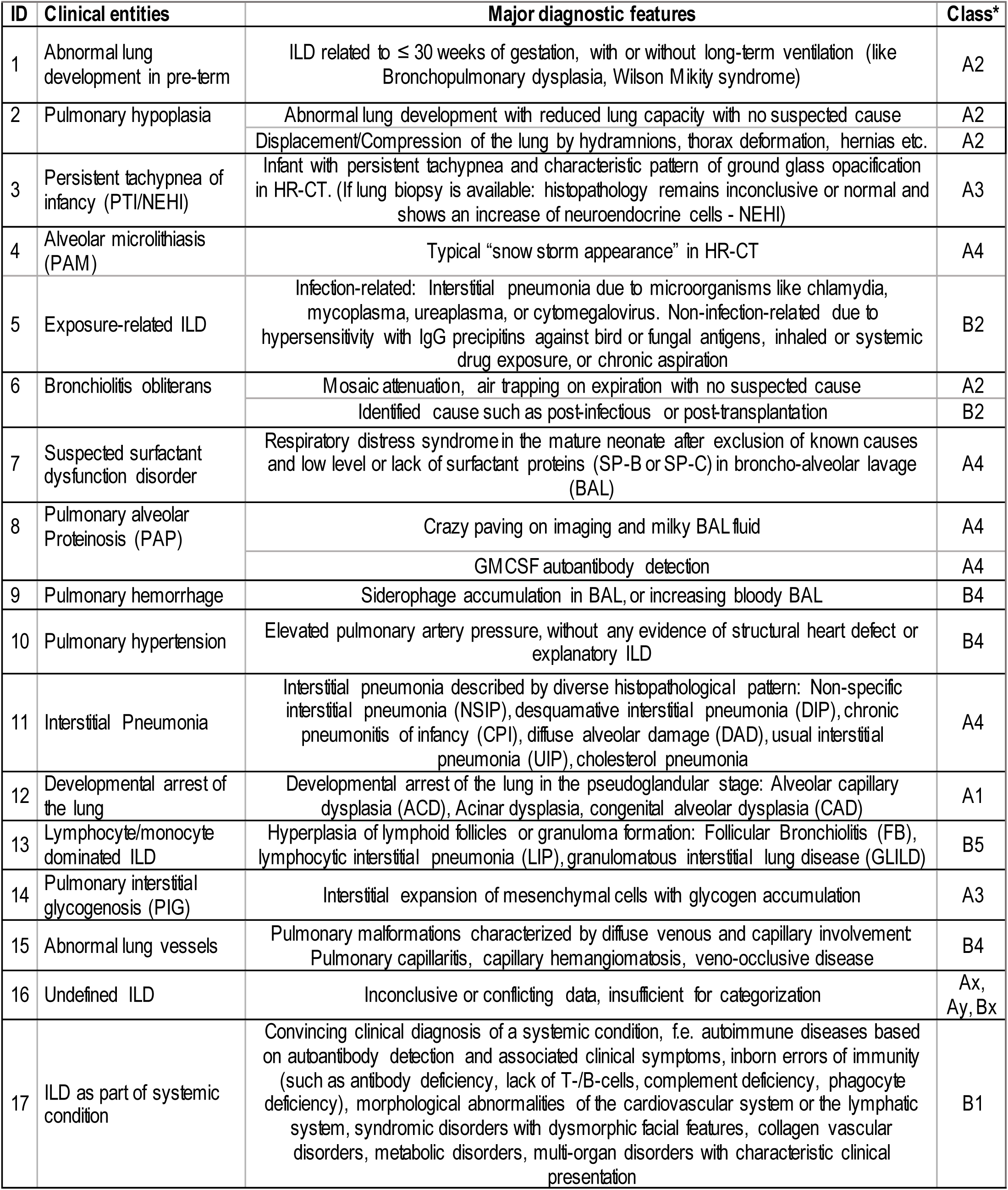
List of clinical entities with their major diagnostic features. A multi-disciplinary team defined differential diagnoses in 1,686 children with ILD. For reference, the allocation of the clinical entities into the current classification of chILD is indicated (Deutsch et al. 2007, Griese et al., 2009). The detailed individual final diagnoses of all patients making up a clinical entity are listed in Tab. S3.

| ID | Clinical entities | Major diagnostic features | Class* |
| --- | --- | --- | --- |
| 1 | Abnormal lung development in pre-term | ILD related to $\leq 30$ weeks of gestation, with or without long-term ventilation (like Bronchopulmonary dysplasia, Wilson Mikity syndrome) | A2 |
| 2 | Pulmonary hypoplasia | Abnormal lung development with reduced lung capacity with no suspected cause | A2 |
|  |  | Displacement/Compression of the lung by hydramnions, thorax deformation, hernias etc. | A2 |
| 3 | Persistent tachypnea of infancy (PTI/NEHI) | Infant with persistent tachypnea and characteristic pattern of ground glass opacification in HR-CT. (If lung biopsy is available: histopathology remains inconclusive or normal and shows an increase of neuroendocrine cells - NEHI) | A3 |
| 4 | Alveolar microlithiasis (PAM) | Typical "snow storm appearance" in HR-CT | A4 |
| 5 | Exposure-related ILD | Infection-related: Interstitial pneumonia due to microorganisms like chlamydia, mycoplasma, ureaplasma, or cytomegalovirus. Non-infection-related due to hypersensitivity with IgG precipitins against bird or fungal antigens, inhaled or systemic drug exposure, or chronic aspiration | B2 |
| 6 | Bronchiolitis obliterans | Mosaic attenuation, air trapping on expiration with no suspected cause | A2 |
|  |  | Identified cause such as post-infectious or post-transplantation | B2 |
| 7 | Suspected surfactant dysfunction disorder | Respiratory distress syndrome in the mature neonate after exclusion of known causes and low level or lack of surfactant proteins (SP-B or SP-C) in broncho-alveolar lavage (BAL) | A4 |
| 8 | Pulmonary alveolar Proteinosis (PAP) | Crazy paving on imaging and milky BAL fluid | A4 |
|  |  | GM-CSF autoantibody detection | A4 |
| 9 | Pulmonary hemorrhage | Siderophage accumulation in BAL, or increasing bloody BAL | B4 |
| 10 | Pulmonary hypertension | Elevated pulmonary artery pressure, without any evidence of structural heart defect or explanatory ILD | B4 |
| 11 | Interstitial Pneumonia | Interstitial pneumonia described by diverse histopathological pattern: Non-specific interstitial pneumonia (NSIP), desquamative interstitial pneumonia (DIP), chronic pneumonitis of infancy (CPI), diffuse alveolar damage (DAD), usual interstitial pneumonia (UIP), cholesterol pneumonia | A4 |
| 12 | Developmental arrest of the lung | Developmental arrest of the lung in the pseudoglandular stage: Alveolar capillary dysplasia (ACD), Acinar dysplasia, congenital alveolar dysplasia (CAD) | A1 |
| 13 | Lymphocyte/monocyte dominated ILD | Hyperplasia of lymphoid follicles or granuloma formation: Follicular Bronchiolitis (FB), lymphocytic interstitial pneumonia (LIP), granulomatous interstitial lung disease (GLILD) | B5 |
| 14 | Pulmonary interstitial glycogenosis (PIG) | Interstitial expansion of mesenchymal cells with glycogen accumulation | A3 |
| 15 | Abnormal lung vessels | Pulmonary malformations characterized by diffuse venous and capillary involvement: Pulmonary capillaritis, capillary hemangiomatosis, veno-occlusive disease | B4 |
| 16 | Undefined ILD | Inconclusive or conflicting data, insufficient for categorization | Ax, Ay, Bx |
| 17 | ILD as part of systemic condition | Convincing clinical diagnosis of a systemic condition, f.e. autoimmune diseases based on autoantibody detection and associated clinical symptoms, inborn errors of immunity (such as antibody deficiency, lack of T-/B-cells, complement deficiency, phagocyte deficiency), morphological abnormalities of the cardiovascular system or the lymphatic system, syndromic disorders with dysmorphic facial features, collagen vascular disorders, metabolic disorders, multi-organ disorders with characteristic clinical presentation | B1 |

### Application of a simple etiological classification approach for chILD

We used each patient’s diagnosis to investigate the applicability of a simple classification approach for chILD, answering three consecutive questions, similar as in every day clinical practice:

#### 1. In which children can we make an etiological diagnosis due to an identified cause/ lung injury?

In 419 children we recognized an ILD with an identified exposure-related cause or lung injury (Fig. 2A,B). These children came from four clinical entities, i.e. abnormal lung development due to extreme prematurity, pulmonary hypoplasia, exposure-related ILD and bronchiolitis obliterans, with their individual final diagnoses and frequencies listed in Tab. S3.

**Fig. 2:**
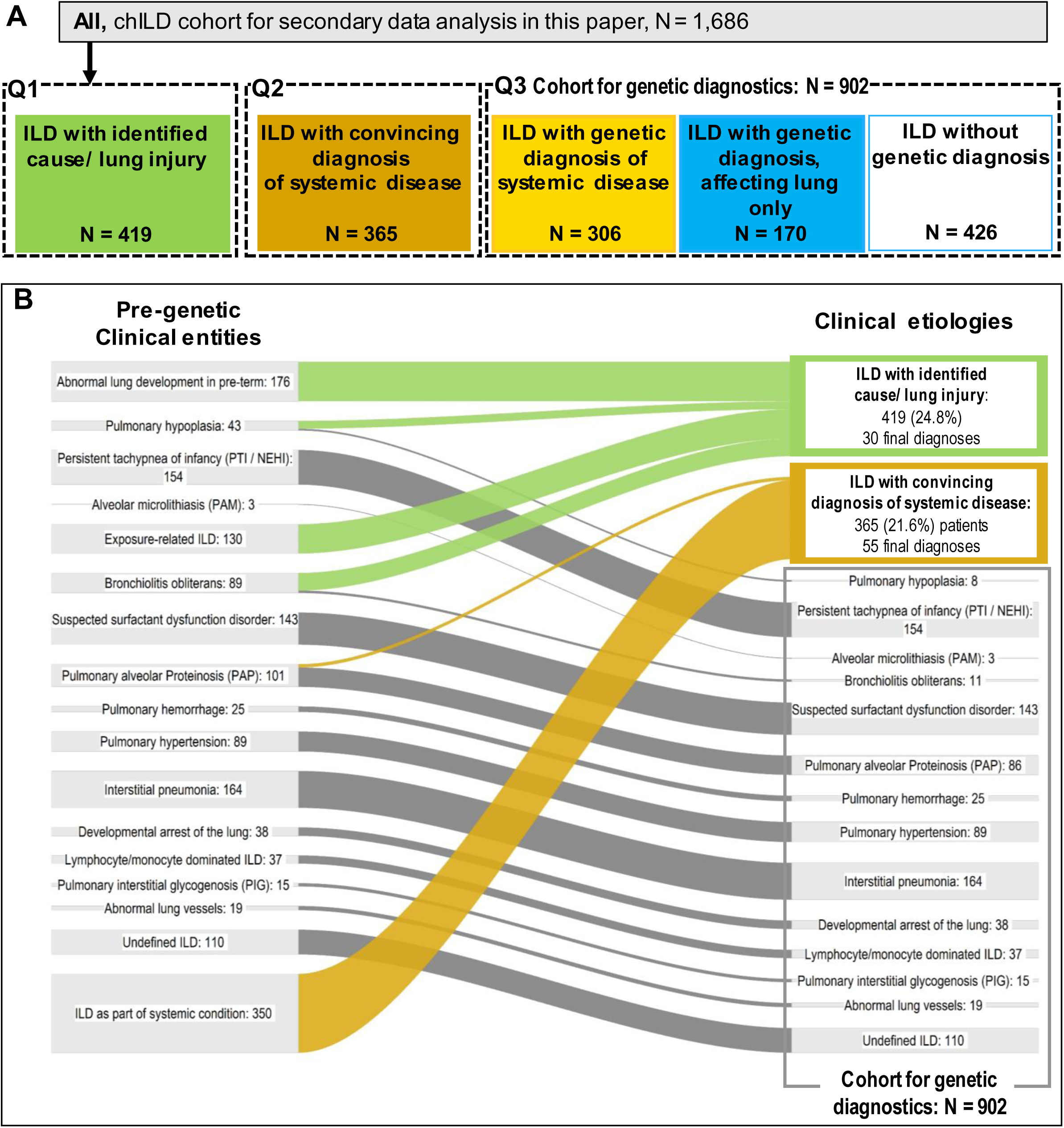
**A: Name and their included number of patients for each aetiological cohort answering hypothesis 1. B:** Each of the 1,686 children initially classified into clinical entities left) were grouped based on their aetiology (right) by answering question 1 and 2, if a exposure-related cause or convincing diagnosis of systemic diseases was able to be identified in children with ILD. Remaining children build the cohort for genetic diagnostics (see Tab. 3)

**Tab. 3:** Genetic investigation of ILD with unclear etiology, N = 902, and their details. Patients are grouped by their clinical entity, given with their total number of patients. All patients underwent genetic analysis. The number of patients with genetic diagnosis is indicated and the identified causal gene listed with their occurrence, if identified more than once. Bold genes were recently discovered or described in the context of this register, including *FARSA*/*FARSB* (2021), *NHLRC2* (2021), *STAT3* (2021), *ZNFX1* (2021), *AGR2* (2022)*, CAPNS1* (2023), *FGF10* (2023), and *CCR2* (2025).

| Clinical entities | N = | Genetic diagnosis, N = (%) | Genetic etiology (details in Tab. S2) |
| --- | --- | --- | --- |
| Pulmonary hypoplasia | 8 | 5 (62.5%) | *EYA1*, ***FGF10***, *HBB*, *TBX4*, *THOC6* |
| Persistent tachypnea of infancy, PTI | 154 | 17 (11.0%) | *SRRM2* (3), *MACF1* (2), *BRWD3*, *CDK13*, *COL4A1*, *DEPDC5*, *IARS1*, *IL6R*, *IL6ST*, *NKX2-1*, *STXBP2*, *TERC*, *TERT*, *UBE3B* |
| Alveolar microlithiasis, PAM | 3 | 3 (100%) | *SLC34A2* (3) |
| Bronchiolitis obliterans | 11 | 6 (54.5%) | ***AGR2*** (2), *CAV1*, *CYBA*, *DOCK8*, *MCM4* |
| Suspected surfactant dysfunction disorder | 143 | 99 (69.2%) | *ABCA3* (52), *SFTPC* (27), *SFTPB* (17), *NKX2-1* (2), *SFTPA1* |
| Pulmonary alveolar Proteinosis, PAP | 86 | 75 (87.2%) | *MARS* (37), *CSF2RA* (19), *SFTPC* (8), *ABCA3* (2), *OAS1* (2), *CSF2RB*, ***FARSA***, *IARS1*, *PNP*, *SLC7A7*, *SFTPB*, *TBX4* |
| Pulmonary hemorrhage | 25 | 9 (36.0%) | *COPA* (4), *TBX4* (3), ***FGF10***, *NLRP3* |
| Pulmonary hypertension | 89 | 50 (56.2%) | *TBX4* (12), *FLNA* (10), *FOXF1* (4), *BMPR2* (2), *CDK13* (2), *ENG* (2), *KCNK4* (2), 11q23 del, 15qdel, 15q11dup, chr19q13dup, 5q trisomy, 7q11del, 9p monosomy, *ACTA2*, *COL4A1*, *COQ2*, *EIF2AK4*, *G6PD*, *KCNK3*, *PIEZO1*, *SLC25A26*, *TMEM173* |
| Interstitial Pneumonia | 164 | 105 (64.0%) | *ABCA3* (39), *SFTPC* (14), *SFTPB* (5), ***FARSB*** (4), ***FARSA*** (3), *COPA* (3), *IFIH1* (3), *NKX2-1* (3), 22q11.2 del (2), 9p Monosomy (2), ***CCR2*** (2), ***STAT3*** (2), *NLRP3* (2), *FLNA* (2), ***ZNFX1*** (2), *TMEM173* (2), *ATM*, *CD40*, *CYBA*, *GATA2*, *HELLS*, *MEFV*, *NCF2*, ***NHLRC2***, *PEX1*, *PLCG2*, *RAB27A*, *TBX4*, *TERC*, *TERT* Xp22.33 dup |
| Developmental arrest of the lung | 38 | 20 (52.6%) | *FOXF1* (16), *STRA6* (2), ***FGF10***, Xp22.33 dup |
| Lymphocyte/monocyte dominated ILD | 37 | 23 (62.1%) | *COPA* (9), ***STAT3*** (2), 22q11.2 del, *CYBA*, *CYBB*, *NKX2-1*, *IL2RG*, *IRF2BP2*, *LRBA*, *SCNN1A*, *SFTPC*, *TERT*, *TNFRSF13B*, *TMEM173* |
| Pulmonary interstitial glycogenosis, PIG | 15 | 1 (6.7%) | *RARB* |
| Abnormal lung vessels | 19 | 4 (21.1%) | *EIF2KA*, ***CAPNS1***, *KDM3B*, X22.33 dup |
| Undefined ILD | 110 | 59 (53.6%) | *TMEM173* (12), *NKX2-1* (12), *FLNA* (3), *CYBB* (2), ***FARSB*** (2), ***STAT3*** (2), ***ZNFX1*** (2), 5q trisomy, Xp22.33 dup, *ADA*, *ACTA2*, *ATM*, *ATR*, *CSF2RA*, *DKC1*, *DOCK8*, *FOXP3*, *HBA2*, *HBB*, *IFIH1*, *IL2RG*, *KDM6A*, *NHLRC2*, *PCDH19*, *PLCG2*, *RTEL1*, *SLC25A20*, *TBX4*, *TERT*, *TSC1*, *WFS1* |
|  | **902** | **476 (52.8%)** |  |

#### 2. In which children can we make the etiological diagnosis of a specific ILD as part of a systemic disease?

In 365 children, the respiratory symptoms were most likely attributable to an underlying systemic condition, often diagnosed prior to pulmonary manifestations (Fig. 2A,B). The latter was reflected in the later onset of respiratory symptoms, at an average age of 5.2 years (p <0.001 compared to other groups, Tab. 1). We attributed all symptoms to a single condition, acknowledging the impossibility to distinguish if symptoms arose from a systemic disease or occur independently. Examples of the selected conditions included sarcoidosis, systemic vasculitis, ILD associated with autoimmune conditions such as autoimmune pulmonary alveolar proteinosis or lupus erythematosus, and rheumatologically conditions with ILD like systemic sclerosis. For the full list with frequencies of all conditions and available histopathological results refer to Tab. S3.

Application of these two questions was able to etiologically recognize 784 of the 1,686 children (Fig. 2B). Among 902 children, no disease etiology was identified, prompting an evaluation of the role of genetic testing.

#### 3. In how many children without an etiological ILD diagnosis based on an identified cause/ lung injury or a systemic condition can we identify a genetic cause and which?

All 902 children had some genetic testing in their ILD work-up (Fig. 3A), which led to the identification of a causal genetic diagnosis in more than half (476 of 902; 52.8%, Tab. 3). The diagnostic yield was different between clinical entities (Fig. 3B, Tab.3). In children with pulmonary alveolar proteinosis, suspected surfactant dysfunction disorders or interstitial pneumonia causal variants were identified in 87.2%, 69.2%, and 64.0%. In about half of the patients with suspected surfactant dysfunction disorder and one third of the patients with interstitial pneumonia, a disease-causing variant in *ABCA3, SFTPC,* or *SFTPB* was identified. These genetic etiologies, along with patients harboring disease-causing variants in *SLC34A2*, were collectively grouped as having ILD with genetic diagnosis affecting lung only. In total, this subgroup comprised 170 patients (Fig. S1, blue fluxes). Beyond those known candidate genes for ILD, in 306 patients various genetic conditions associated with systemic diseases were identified (Tab. 3, Fig. S1, yellow fluxes).

**Fig. 3:**
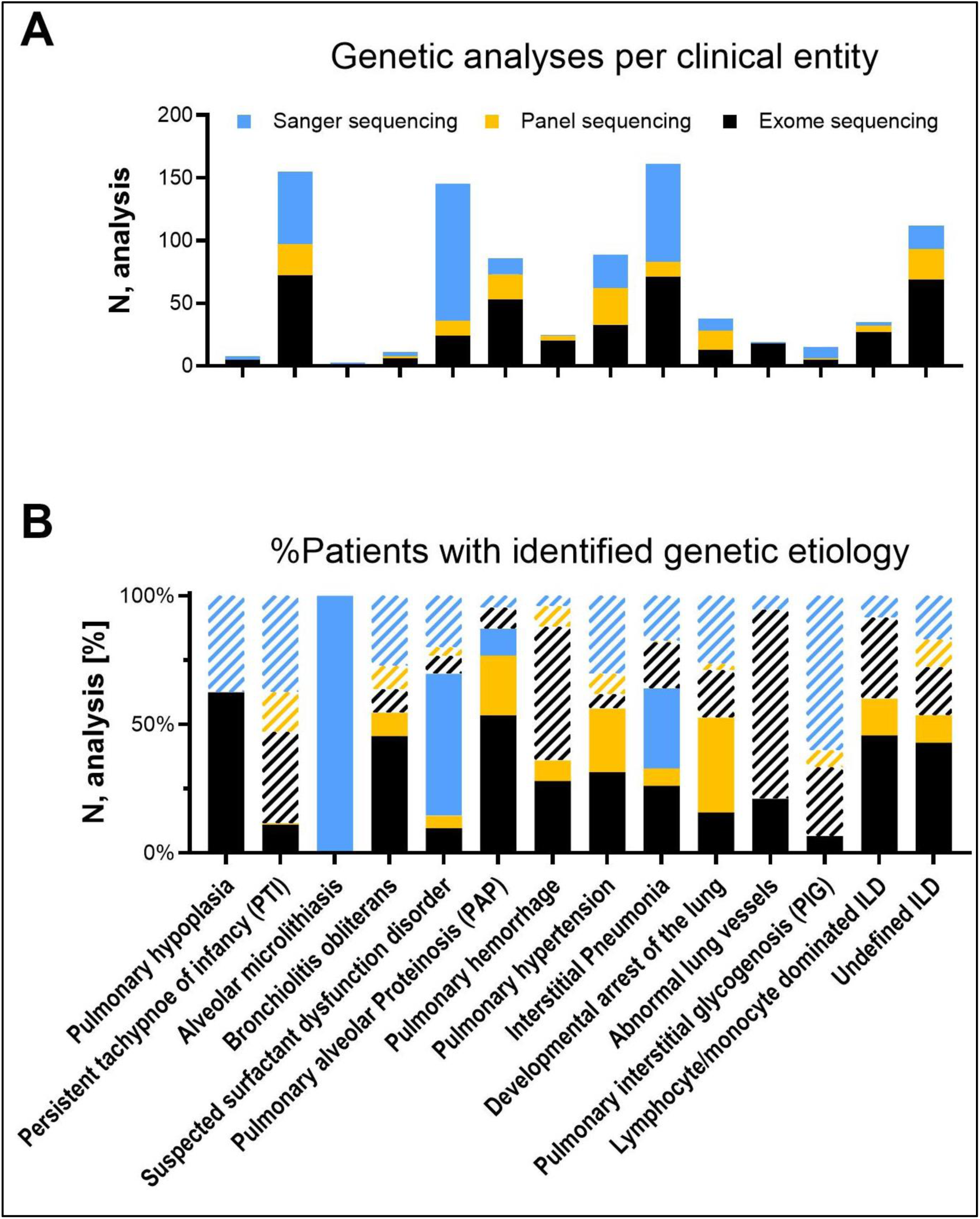
**A: Different type of genetic testing in total numbers in patients with ILD of unknown aetiology (N = 902) and B) their success rate, showing if genetic aetiology was identified (N = 476) or not (N = 426; white stripes).** Sanger in blue, panel sequencing in yellow and exome sequencing in black.

In 426 patients, no causal genetic variant could be identified. These patients predominantly originated from clinical entities for which either no genetic cause has yet been established or where a monogenetic etiology is considered unlikely, such as PTI/ NEHI or pulmonary interstitial glycogenosis. These conditions represented the lowest diagnostic yield at 11.0% and 6.7%, respectively (Tab. 3).

In summary, among the 1,686 cases, 24.9% were categorized as ILD with an identified cause or lung injury, 21.6% as ILD with a convincing diagnosis of systemic disease, 18.1% as ILD with a genetic diagnosis of systemic disease, 10.1% as ILD with genetic diagnosis affecting the lungs only, and 25.3% as ILD without genetic diagnosis (Fig. 2A). Collectively, an etiological diagnosis was established in 74.7% of the patients, encompassing 179 distinct etiological chILD diagnoses, including 30 exposure-related conditions, 55 diagnoses defined as convincing systemic conditions and 94 genetic diagnoses, including 89 systemic diseases (Tab. S3).

Among these 89 systemic diseases, 25 conditions were linked to genes not yet associated with specific ILDs (Tab. S3). Among these, 11 patients exhibited immune-related disorders linked to *IFIH1* (four patients), *NLRP3* (three patients), *ATR, CD40, HELLS,* and *IRF2BP2*. Additionally, 13 patients had neurological developmental disorders caused by pathogenic variants in *SRRM2* (three patients), *CDK13* (three patients), *MACF1* (two patients), *BRWD3*, *DEPCD5*, *PCDH19*, *STXBP2*, and *UBE3B,* of interest with an over-representation among individuals clinically diagnosed as having PTI/NEHI. Furthermore, a complex multisystemic disorder was identified in 18 patients with pulmonary hypertension observed as primary feature in 39%. While 12 exhibited diverse copy number variations (Xp22.33 duplication in four patients, 9p Monosomy in three patients, 5q Trisomy in two patients, 15q11 duplication, 15q26 deletion, and 19q13 duplication), six patients harbored pathogenic variants in *KCNK4* (two patients), *KDM6A*, *WFS1*, *COL4A1*, and *IARS1*.

### Occasional genetic testing among children with an ILD with identified cause/ lung injury or a convincing diagnosis of systemic disease

Notably, among 784 patients with an etiological chILD diagnosis, i.e. those two groups generated in response to question Q1 and Q2, genetic testing was occasionally performed, which was usually requested by the attending physician to confirm the diagnosis, to formally exclude alternative conditions, or to refine differential diagnoses (Tab. S3).

In the group of 419 children with ILD with an identified cause/ lung injury, genetic testing was performed in 153 cases. Among these, eight children (5.2%) were unexpectedly found to have a systemic condition. Although no linkage described, some of these conditions might be related to the ILD (see Tab. S4, clinical entities 7 and 9), while others were likely unrelated (Tab. S4, clinical entities 1 and 4). Importantly, none of these children had genetic variants in well-known childhood ILDs, such as surfactant dysfunction disorders. In the next group of children with ILD with convincing diagnosis of a systemic disease, genetic testing was performed in 158 cases. In all of these, the genetic results confirmed the clinical diagnosis (Tab. S4). These findings show that broad genetic testing can help uncover previously undiagnosed systemic conditions. However, they also demonstrate that limited testing for surfactant-related genetic variants is not required when ILD is caused by a known injury or condition.

### Non-pulmonary organ involvement increases the likelihood of an underlying systemic disease etiology for ILD two to three-fold

We systematically recorded the non-pulmonary organ involvement in all 1,686 chILD patients to evaluate their role for the etiological diagnosis of ILD. Patients with ILD as part of a systemic condition (genetic or clinical diagnosis) had approximately three-fold more HPO-terms and two-fold more organs involved (Fig. 4A,B). Overall organ involvement was then grouped into organ-specific comorbidities and ranked by their occurrence, showing vascular, cardiac, neurological, and gastrointestinal organs as most commonly affected in all children with ILD (Fig. 4C,D). ILD associated with a convincing diagnosis of systemic disease, and to a slightly lesser extent, patients with a genetic diagnosis of systemic disease, demonstrated more frequent involvement across all organs (Fig. 4C). We next stratified patients distinguishing between those with ILD as part of a systemic condition (‘ILD, systemic’), i.e. either by clinical or genetic diagnosis, and those in whom ILD presented as an isolated lung condition (‘ILD, lung only’), attributed to a defined cause/ lung injury or a genetic diagnosis affecting lung only. Notably, over 65% of patients grouped as ‘ILD, lung only’ had no or only one additional organ involvement (Fig. 5A). When two or more organs were affected in a patient with ILD, the likelihood of an underlying systemic disease surpassed 50%, further increasing as the number of involved organs rose (Fig. 5B). Given the non-negligible frequency of non-pulmonary organ involvement in general (Tab. S5), the impact of specific organ systems was assessed. The presence of autoimmune, lymphatic, immunodeficiency, endocrine, vascular, dermal, or syndromic manifestations was associated with a significantly increased probability of an underlying systemic condition (Fig. 5C, 95%CI>1). In contrast, neurological, nephrotic, and skeletal abnormalities were prevalent in both subgroups with no predictive differentiation.

**Fig. 4:**
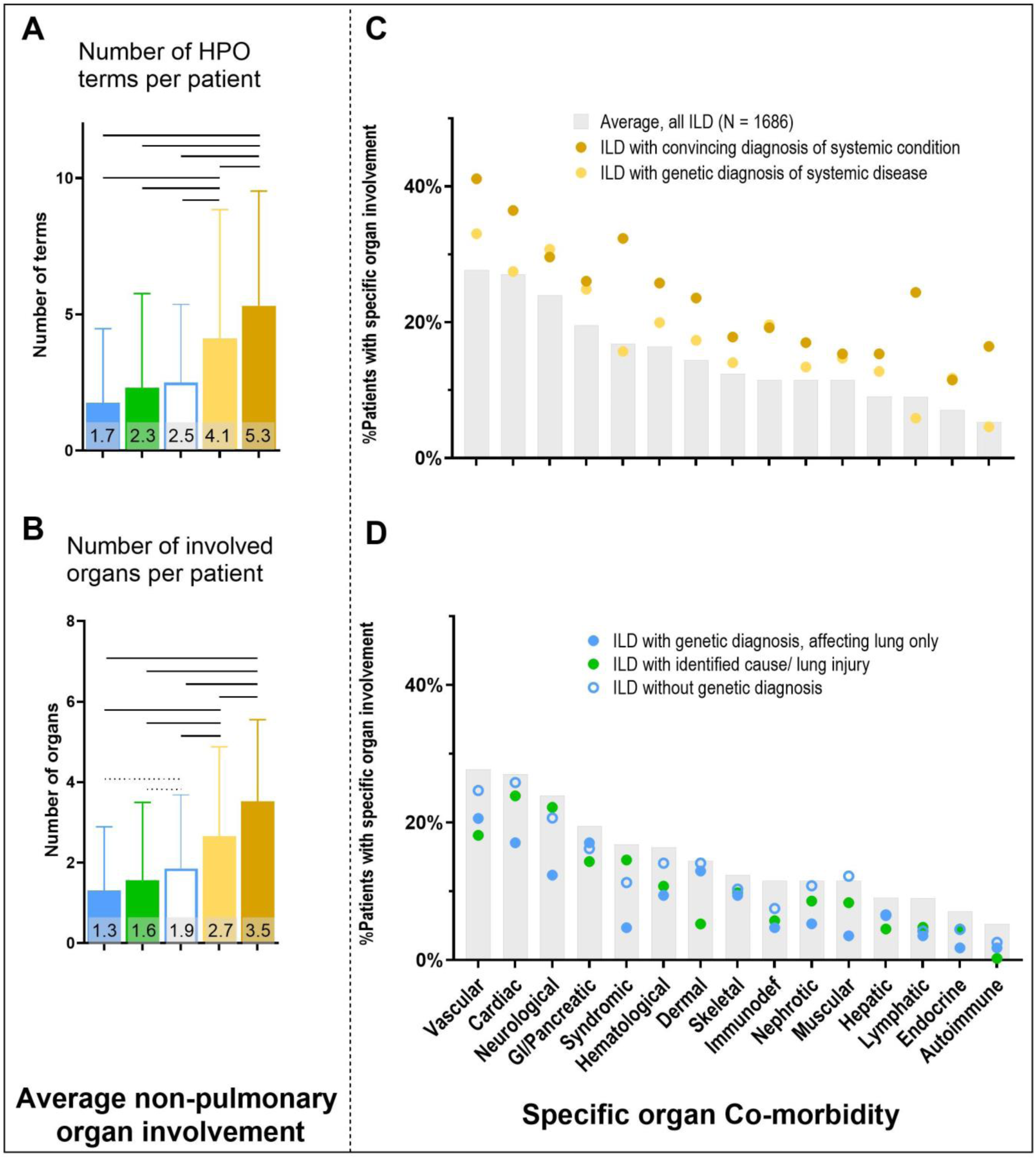
Non-pulmonary organ involvement, in the five aetiological cohorts: ILD with genetic diagnosis affecting lung only (blue), ILD with identified cause/ lung injury (green), ILD without genetic diagnosis (blue outline), ILD with genetic diagnosis of systemic disease (bright yellow), or ILD with convincing diagnosis of systemic disease (dark yellow). **A: Average Number of HPO terms per patient** describing non-pulmonary organ involvement and **(B) the average number of individual organs involved per patient** (Data are given as mean ±SD, significance is depicted as solid line: p<0.0005: **** and dotted line: p <0.05: *). **C, D: Differentiation of organ involvement into the organ-specific co-morbidity.** All patients (N = 1,686) grey columns, patients with underlying systemic diseases and patients with ILD with identified cause/ lung injury, ILD with genetic diagnosis affecting the lung only and ILD without genetic diagnosis.

**Fig. 5:**
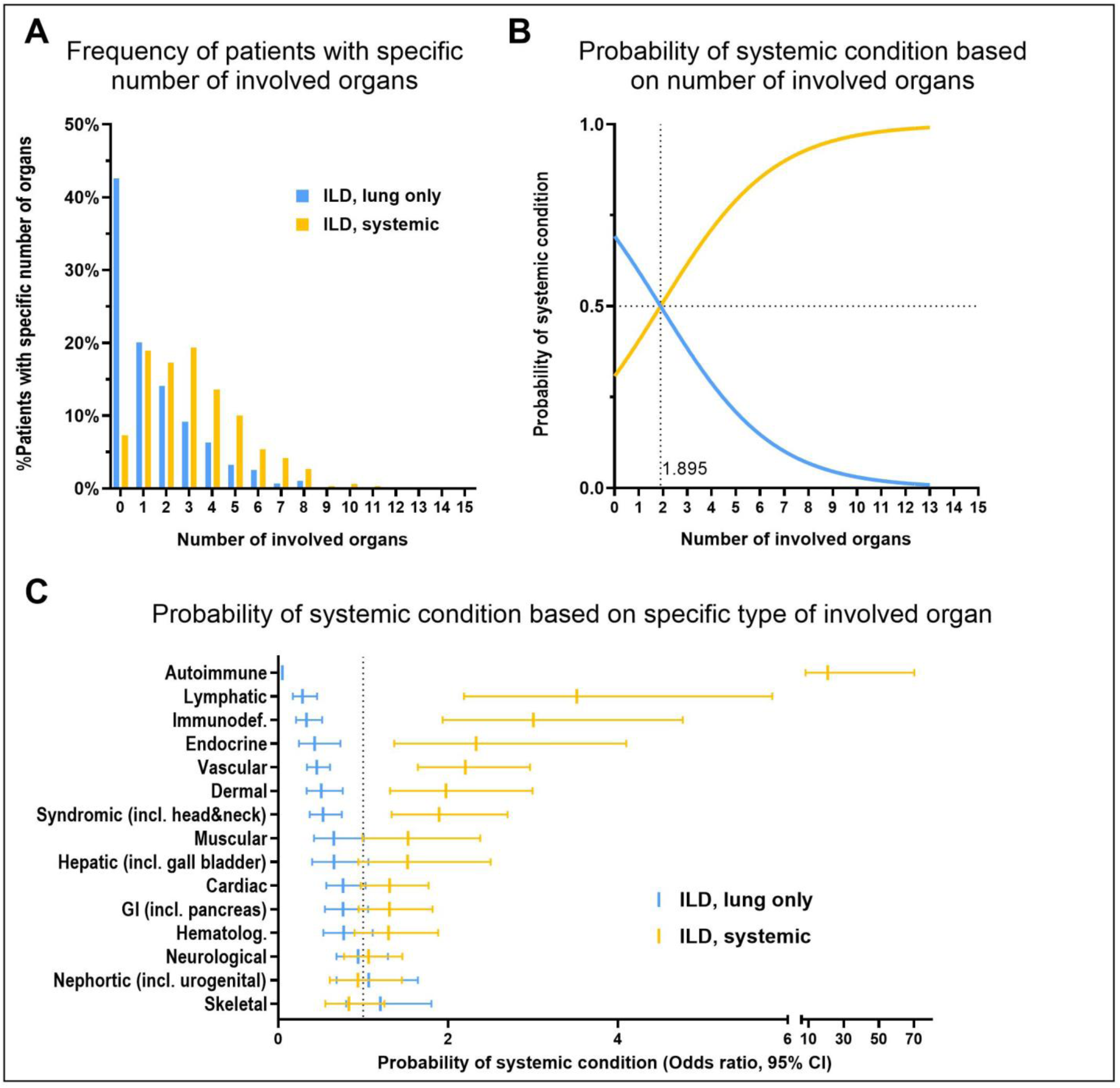
Probability of an underlying systemic disease in children with ILD. A: Frequency of patients with one to fifteen involved organs in patients with an isolated lung disease (in blue: patients with identified cause/lung injury and patients with genetic diagnosis of ILD; “ILD, lung only”) compared to patients with ILD as part of an underlying systemic condition (in yellow: clinical or genetic diagnosis of systemic disease; “ILD, systemic”). B: Probability of an underlying systemic condition based on the number of affected organs. C: Results of multiple logistic regression given the observed probability of a systemic condition depending on the type of affected organs, data is given as odds ratio with 95% CI.

## DISCUSSION

This study applied a practical, stepwise diagnostic classification to a large cohort of 1,686 chILD patients, all previously diagnosed by a multidisciplinary team. The approach enabled clear etiological grouping based on simple, well-defined criteria and demonstrated several advantages.

### Etiology-based grouping can help guiding patient care

Etiology-based grouping supports clinical decision-making. In children with an etiological diagnosis due to an identified exposure-related cause, eliminating ongoing exposures is critical. A wait-and-see approach may be appropriate in cases of past lung injuries resulting from lung immaturity or displacement/compression. If disease progression is unexpectedly severe, recurrent infections suggest an underlying immunologic disorder, or recurrent aspiration raises concern for a neurological or syndromic condition, exome or genome sequencing should be re-considered. In children with an ILD due to a systemic disease, multidisciplinary collaboration involving non-pulmonary subspecialties may be required for comprehensive patient management. Identifying systemic diseases underlying chILD enhances prognostic accuracy and enables rational selection of targeted, personalized treatment strategies and monitoring for disease-related complications before they occur. Even as genetic testing becomes more accessible and affordable, we believe that broad genetic testing is not justified in all suspected chILD cases. Despite its benefits, advanced genetic testing presents challenges, particularly the need for competent genetic consulting to ensure patients and families receive accurate interpretation, informed decision-making, and appropriate clinical guidance(21, 22). Lastly the resource-intensive nature, including time for interpretation and data storage, need to be considered.

### High yield of etiology-based genetic diagnostics in chILD

In a well-defined cohort of etiologically undiagnosed chILD, a causal genetic diagnosis was achieved in 54% of cases (476 of 902). Within specific clinical entities such as suspected surfactant dysfunction disorders or pulmonary alveolar proteinosis, the diagnostic yield ranged from 69% to 87%. Among 902 genetically analyzed patients, targeted sequencing was performed in 678, identifying a genetic cause in 253. Among the 649 unsolved patients, including 224 without prior genetic testing, 415 underwent exome sequencing, identifying a genetic cause in 223. The other 234 patients were evaluated solely by targeted sequencing, which remained inconclusive. In these cases, exome sequencing may further close the diagnostic gap.

Importantly, even when genetic analysis is inconclusive, exome data allow for future reinterpretation as genomic knowledge evolves. This long-term utility reinforces the value of early integration of genetic testing into clinical workflows. Our data support the view that genetic testing may reduce the need for lung biopsies. The number of lung biopsies declined steadily over 5-year periods, particularly after the widespread adoption of exome sequencing in 2019: from 81 cases in 2010-2014, to 79 in 2015-2019, and only 49 in 2020-2024. Simultaneously, the number of genetic tests performed increased from 159 over 353 to 439, respectively.

### Flexible adaptation to diagnostic advances

Not linking the classification to specific diagnostic tests, such as lung biopsies(1), avoids creating rigid hierarchies and allows for the seamless integration of newly discovered diseases and emerging genomic insights under common clinical entities. This flexibility is critical in a rapidly evolving diagnostic landscape.

In this context, genetic analysis confirmed known diagnoses and also facilitated the discovery of potential novel candidate genes. In our study, 42 patients harbored pathogenic variants in 25 genes not previously linked to an ILD phenotype (Tab. S3). Although definitive proof of causality remains elusive, recurrence of variants in three to four unrelated individuals suggests a possible connection. Support for these associations may stem from treatment response, functional studies in cell or animal models(20), or the accumulation of similar cases. Therefore, it is important to carefully document and report these observations.

### Role of non-pulmonary organ involvement in patient management

In a large chILD cohort we determined the prevalence of 179 distinct etiological chILD diagnoses, of which approximately 40% were systemic conditions. We also quantified for the first time the extent of non-pulmonary organ involvement in chILD, demonstrating that the presence of ≥two non-pulmonary organ manifestations increased the likelihood of an underlying systemic disease by two to three-fold. Remarkably, approximately 15% (47 of 306) of patients with a genetic diagnosis of systemic disease exhibited no additional organ manifestation. This finding is clinically significant, as it suggests that certain characteristic symptoms may be subtle, overlooked, or emerge later during the disease course. Beyond the number of affected organs, particularly when more than two are involved, the type of organ should raise the suspicion for systemic diseases. Consequently, exome sequencing maybe considered if ≥two non-pulmonary organs are involved, the initial diagnosis made does not fit the expected disease course or a second underlying condition is suspected. Taking the etiology-based classification, which was successfully applied here, into clinical practice may help manage the broad spectrum of chILD.

## Data availability statement

Anonymized aggregated data from the chILD-EU register are available upon reasonable request. Research proposals for such data can be submitted to the corresponding author and will be assessed by the chILD-EU register coordination team. Aggregated data will be shared if the proposal addresses relevant research questions. Requests for anonymized individual patient data should also be directed to the corresponding author, who will facilitate contact with the responsible physicians to discuss the legal and administrative feasibility of data sharing.

## Acknowledgments

We thank all parents and children for participating in the study and researchers, technicians, pediatricians who helped with data collection.

## chILD-EU register contributors

*Austria* A Pfleger, E Eber (Graz), A Zschocke (Innsbruck), K Kainz, Z Szepfalusi, A Zacharasiewicz (Vienna). *Belgium* M Proesmans (Leuven), H Schaballie (Ghent). *Brazil* LV Ribeiro Silva Filho (São Paulo). *Czech Republic* V Koucký (Prague). *Denmark* F Buchvald, AM Ring (Copenhagen), S Rubak (Aarhus). G*ermany* P Aleksander, S Lau, A Lejeune, J Röhmel, M Stahl, L Tetteh (Berlin), S Dillenhoefer, C Koerner-Rettberg, A Schlegtendal, A Wiemers, N Teig (Bochum), L Lange (Bonn), P Kaiser-Labusch (Bremen), S Becker (Darmstadt), C Vogelberg (Dresden), D Schramm, A Schuster (Dusseldorf), F Stehling, M Steindor (Essen), J Trischer (Frankfurt), C Müller (Freiburg), A Bagheri - Potthoff, L Nährlich (Giessen), R Scheidmann (Hamburg), P Maier, J Carlens, D Renz, N Schwerk, M Wetzke, AH Ola (Hanover), L Eberhard, S Hämmerling, A Krümpelmann, O Sommerburg (Heidelberg), D Schöndorf (Homburg), R Körner, E Rietschel, J Schönenkorb, J Thomassen (Cologne), F Prenzel (Leipzig), L Sultansei (Luebeck), D Schreiner (Mainz), S Bergemann, M Forstner, A Gold, F Gothe, B Kammer, M Kappler, C Klein, C Kröner, I Krueger-Stollfuss, J Ley-Zaporozhan, K Michel, E Reisch, K Reiter, M Rohlfs, J Rodler, A Schams, N Tran (Munich), J Große-Onnebrink (Muenster), A Kiefer, E Plattner (Regensburg), N Baumann, M Rose, J Papenkort (Stuttgart), W Baden (Tuebingen), S Bode, H Ehrhardt (Ulm), S Reu-Hofer, J Sammler (Wuerzburg). *Greece* E Manali (Athens). *Italy* R Cutrera, N Ullmann (Roma), A Baritussio (Padua). *Poland* K Krenke, J Lange, H Marczak (Warsaw). *Portugal* T Bandeira (Lisbon). *Spain* A Moreno-Galdó, A Torrent-Vernetta (Barcelona), S Castillo-Corullon, J Lopez, S Pérez-Tarazona (Valencia). *Switzerland* M Hitzler, E Seidl, (Zurich), I Rochat (Lausanne), N Regamey (Lucerne). *Turkey* G Cinel, N Coganoglu, N Emiralioglu, N Kiper, T Sysmanlar (Ankara), AA Klinic (Istanbul). *United Kingdom* S Cunningham, M MacLean (Edinburgh), S Mayell (Liverpool), A Bush, I Lancoma-Malcolm, C Nwokoro (London), J Bhatt, C Youle (Nottingham)

