## Supplement figures and tables for "Childhood interstitial lung disease – application of a stepwise diagnostic classification"

Christina K Rapp[1], Matthias Griese[1] for the chILD-EU consortium core (Nicolaus Schwerk, Julia Carlens, Florian Stehling, Julia Rodler, Honorata Marczak, Katarzyna Krenke) and contributor† group

† Contributors to the chILD-EU register are listed at the end of the article
1 Dr. von Haunersches Kinderspital, University of Munich, German Center for Lung Research, Munich, Germany

**Tab. S1: Definition of five etiological groups**

| | | |
|---|---|---|
| Q1 | ILD with identified cause/ lung injury | Identified (exposure-related) causes or lung injuries including lung prematurity from birth below 30 weeks of gestation and significant respiratory impairment, pulmonary hypoplasia due to lung compression/ displacement, and interstitial pneumonia caused by pathogens like Mycoplasma or non-infection causes due to IgG-mediated hypersensitivity to antigens, inhaled or systemic drug exposures, or chronic aspiration |
| Q2 | ILD with convincing diagnosis of a systemic disease | Individuals presenting with a characteristic clinical phenotype consistent with a well-defined condition plausibly accounting for observed ILD manifestations. The term "convincing" reflects the diagnostic likelihood and includes definite diagnoses (probability >90%) |
| Q3 | ILD with genetic diagnosis, affecting lungs only | Genetic variant classified as disease-causing in a gene exclusively associated to ILD such as ABCA3, SFTPC, SFTPB, and SLC34A2 |
| Q3 | ILD with genetic diagnosis of a systemic disease | Genetic variant classified as disease-causing in a gene associated with a systemic condition |
| Q3 | ILD without genetic diagnosis | Genetic analysis was inconclusive or negative |

**Tab. S2: Demographic data of chILD patients included in secondary data analysis. Data were given as total number and percentages per group or median with first and third quartiles.**

| | **chILD-EU register patients** | **Munich centre patients** | **All patients** |
|---|---|---|---|
| Patients with insufficient data, N | 117 | 89 | 206 |
| Patients with secondary immunodeficiency, N | 23 | 86 | 109 |
| **Patients for secondary data analysis, N** | **782** | **904** | **1686** |
| Age at disease onset [years] | 2.4 (0.5 to 8.0) | 0.8 (0.2 to 5.4) | 1.3 (0.3 to 8.0) |
| Patients below the age of 2, N (%) | 364 (46.5%) | 582 (64.4%) | 946 (56.1%) |
| Follow-up [years] | 6.1 (3.0 to 8.5) | 9.8 (2.9 to 15.8) | 7.1 (3.0 to 13.2) |
| Male to female ratio | 1 : 0.89 | 1 : 0.77 | 1 : 0.82 |
| Age at death/lung transplantation [years] | 0.9 (0.2 to 5.3) | 0.4 (0.1 to 1.3) | 0.5 (0.1 to 3.4) |
| Deceased/lung transplanted, N (%) | 133 (17.0%) | 190 (21.0%) | 323 (19.2%) |
| European | 774 | 881 | 1655 |
| Non-European | 8 | 23 | 31 |

**Tab. S3: Children with confirmed ILD (N = 1,686) used for this secondary data analysis. For each patient the diagnosis and diagnostic tests done, were allocated to a clinical entity and the patients were grouped into the most likely etiological group**: ILD with identified cause/ lung injury (green, N = 419) and ILD with convincing diagnosis of a systemic condition (N = 365, dark yellow). For the remaining children results of genetic testing were considered and they were grouped into: ILD with genetic diagnosis of a systemic disease (yellow, N = 306), ILD with genetic diagnosis (blue, N = 170) and ILD without genetic diagnosis (white, N = 426). Final diagnosis with major pattern in histopathology if lung biopsy was available (with their occurrence, if identified more than once) and total number of patients given. Alveolar capillary dysplasia (ACD), bronchopulmonary dysplasia (BPD), cholesterol pneumonia (CP), chronic pneumonitis of infancy (CPI), congenital alveolar dysplasia (CAD), desquamative interstitial pneumonia (DIP), diffuse alveolar damage (DAD), diffuse lymphoid hyperplasia (DLH), follicular bronchiolitis (FB), granulomatous interstitial lung disease (GLILD), lymphocyte interstitial pneumonia (LIP), neuroendocrine cell hyperplasia (NEHI), no diagnosis (non-dx), non-specific interstitial pneumonia (NSIP), pulmonary alveolar proteinosis (PAP), pulmonary capillary hemangiomatosis (PCH), pulmonary hypertension (PH), pulmonary interstitial glycogenosis (PIG), pulmonary veno-occlusive disease (PVOD), usual interstitial pneumonitis, (UIP). ***Association between chILD and systemic disease has not yet been described.

| Clinical Entity | | Final diagnosis | Major histological pattern (N), in available lung biopsies | N |
|---|---|---|---|---|
| | **ILD with identified cause/ lung injury** | | | **419** |
| 1 | Abnormal lung development in pre-term | Bronchopulmonary dysplasia in preterm (≤29 weeks of gestation) | Alveolar hypoplasia (11), BPD (9), chronic bronchiolitis (2), DAD (2), fibrosis (2), NSIP (2), PH (2), aspiration pneumonia, emphysema, non-dx, PIG | 160 |
| | | Pulmonary interstitial emphysema (ventilation-induced) | - | 3 |
| | | Wilson Mikity, new BPD | Alveolar hypoplasia | 13 |
| 2 | Pulmonary hypoplasia | Pulmonary hypoplasia associated with diaphragmatic hernia | Alveolar hypoplasia (3), pulmonary hypoplasia, PIG | 12 |
| | | Pulmonary hypoplasia associated with fetofetal transfusion syndrome | - | 1 |
| | | Pulmonary hypoplasia associated with hiatus hernia | Pulmonary hypoplasia | 1 |
| | | Pulmonary hypoplasia associated with hydrothorax | Pulmonary hypoplasia, Pulmonary hypoplasia with PH | 6 |
| | | Pulmonary hypoplasia associated with neuromuscular dysfunction | - | 1 |
| | | Pulmonary hypoplasia associated with oligohydramnios | Alveolar hypoplasia, Pulmonary hypoplasia with fibrosis | 5 |
| | | Pulmonary hypoplasia associated with omphalozele | - | 2 |
| | | Pulmonary hypoplasia associated with polyhydramnios | - | 3 |
| | | Pulmonary hypoplasia associated with thoracic dystrophies | Pulmonary hypoplasia | 4 |
| 5 | Exposure-related ILD | Aspiration syndrome | Aspiration pneumonia (2), chronic bronchiolitis, constrictive bronchitis | 35 |
| | | BO, toxic inhalation | Obliterative bronchiolitis | 2 |
| | | Chlamydia pneumonia | - | 3 |
| | | Cytomegalovirus (CMV) interstitial pneumonia | Infection | 1 |
| | | Diffuse panbronchiolitis | - | 3 |
| | | Drug-related lung injury | - | 3 |
| | | Exogen allergic alveolitis/ hypersensitivity pneumonitis | Hypersensitive pneumonitis (14), NSIP (2), chronic bronchiolitis, GLILD, UIP | 61 |

| | | | | |
|---|---|---|---|---|
| | | Gestational diabetes induced lung underdevelopment and delayed maturation | PIG | 4 |
| | | Intrauterine growth retardation due to maternal alcohol consume | - | 1 |
| | | Mac-Leod-Swyer-James syndrome | Obliterative bronchiolitis, NSIP | 5 |
| | | Meconium aspiration | - | 5 |
| | | Mycoplasma pneumonia | - | 1 |
| | | Pneumocystis jerovecii pneumonia (PJP) | Alveolar hypoplasia | 7 |
| | | Radiation-related lung injury | NSIP | 2 |
| | | Stevens-Johnson syndrome | - | 1 |
| | | Ureaplasma induced interstitial pneumonitis | - | 2 |
| 6 | Bronchiolitis obliterans | BO, post-infectious | Chronic bronchiolitis (9), obliterative bronchiolitis (8), constrictive bronchiolitis (5), bronchitis plastica (2), Infection (3), Non-dx (3), NSIP | 72 |
| | **ILD with convincing diagnosis of systemic disease** | | | **365** |
| 8 | Pulmonary alveolar Proteinosis | Autoimmune PAP | PAP | 15 |
| 17 | ILD as part of systemic condition | Antibasement membrane antibody disease (Goodpasture's syndrome) | Obliterative bronchiolitis | 3 |
| | | Antibody deficiency associated ILDs | Organising pneumonia (2), chronic bronchitis, LIP | 7 |
| | | Antisynthetase syndrome | FB | 3 |
| | | Atypical hemolytic uremic syndrome | Thromboembolic | 1 |
| | | Behcet's disease | - | 2 |
| | | Birt-Hogg-Dube syndrome | Chronic bronchiolitis | 1 |
| | | Cakuthed syndrome | - | 1 |
| | | Cantu syndrome | Alveolar hypoplasia, NSIP | 3 |
| | | Cat eye syndrome | - | 4 |
| | | Charge syndrome | - | 1 |
| | | COFS syndrome (Cerebrooculofacioskeletal syndrome) | - | 1 |
| | | Collagen vascular disorders | Haemosiderosis, chronic bronchiolitis, obliterative bronchiolitis, GLILD | 9 |
| | | Congenital heart disease/ cardiac dysfunction | PIG (8), alveolar hypoplasia, haemosiderosis, PH, Fibrosis (2) | 33 |
| | | Cornelia-de-Lange syndrome | - | 1 |
| | | DiGeorge syndrome | Haemosiderosis, non-dx | 5 |
| | | Down syndrome | Alveolar hypoplasia (7), PH (3), aspiration pneumonitis (2), DAD (2), chronic bronchiolitis, DIP, FB, GLILD, lymphangiectasis, NSIP | 58 |
| | | Ehlers Danlos | - | 2 |
| | | Eosinophilic granulomatosis with polyangiitis (EGPA, Churg Strauss) | Eosinophilic pneumonitis (2), haemosiderosis (2), non-dx | 9 |
| | | Gaucher disease | - | 1 |
| | | Goldenhaar syndrome | - | 2 |
| | | Granulomatosis with polyangiitis (GPA, Wegener) | GLILD (4), chronic bronchiolitis (2), non-dx (2), haemosiderosis | 17 |
| | | Hemoglobinopathy | - | 2 |
| | | Haemorrhagic telangiectasia/ diffuse arteriovenous malformations | Haemosiderosis (2), PH (2), chronic bronchiolitis | 11 |

| | | | | |
|---|---|---|---|---|
| | | Hepatopulmonary syndrome | NSIP, lymphangiectasis, non-dx | 4 |
| | | Hermansky Pudlak syndrome | NSIP, DLH, PH, non-dx | 10 |
| | | Hunter syndrome | PIG | 1 |
| | | IgA vasculitis (Schoenlein-Hennoch) | - | 1 |
| | | Immune dysregulation/ immunodeficiency associated ILD | CP, NSIP, LIP | 19 |
| | | Johanson-Blizzard syndrome | - | 1 |
| | | Kidney, lung, skin disease | Alveolar hypoplasia | 2 |
| | | Klinefelter syndrome | Alveolar hypoplasia | 3 |
| | | Lane Hamilton syndrome | Haemosiderosis (2), FB | 9 |
| | | Langerhans cell histiocytosis | Langerhans cell histiocytosis (7) | 7 |
| | | Lymphangiectasis with/without chylothorax | Lymphangiectasis (12), PIG | 14 |
| | | Lymphangiomatosis | Lymphangiomatosis (3) | 3 |
| | | Lysinuric proteinuria | PAP | 6 |
| | | Marfan syndrome | Emphysema, LIP | 3 |
| | | Microscopic polyangiitis | Chronic bronchiolitis | 2 |
| | | Mixed connective tissue disease | LIP | 6 |
| | | Mucolipidosis type II | - | 1 |
| | | Musculoskeletal disorder associated ILD | NSIP | 5 |
| | | Neurological disorder associated ILD | NSIP (2), chronic bronchiolitis, CP | 16 |
| | | Niemann-Pick associated ILD | PAP (3) | 9 |
| | | Noonan syndrome | NSIP, fibrosis, pulmonary hypoplasia | 8 |
| | | Phagocyte deficiency associated ILD | Cryptogenic bronchiolitis, non-dx | 2 |
| | | Psoriastic arthrithis associated ILD | - | 1 |
| | | Sanfilippo syndrome | PIG | 3 |
| | | Sarcoidosis | Sarcoidosis (8), GLILD, non-dx | 19 |
| | | Systemic autoinflammation associated ILD | - | 2 |
| | | Systemic juvenile idiopathic arthritis | NSIP, PAP, PH, non-dx | 5 |
| | | Systemic lupus erythematosus | DLH, NSIP | 6 |
| | | Systemic sclerosis associated ILD | - | 3 |
| | | Tricho-hepato-enteric syndrome | - | 1 |
| | | Wolf-Hirschhorn syndrome | - | 1 |
| **ILD with genetic diagnosis of systemic disease** | | | | **306** |
| 2 | Pulmonary hypoplasia | Beaulieu-Boycott-Innes syndrome, THOC6 | Alveolar hypoplasia | 1 |
| | | Branchiootorenal syndrome, EYA1 | - | 1 |
| | | DiGeorge syndrome, chr22q11.2 deletion | Alveolar hypoplasia | 1 |
| | | Lacrimo-auriculo-dental-digital syndrome, FGF10 | Alveolar hypoplasia | 1 |
| | | PAH with small patella syndrome, TBX4 | Pulmonary hypoplasia | 1 |
| 3 | Persistent tachypnea of infancy (PTI/NEHI) | Brain small vessel disease, COL4A1* | - | 1 |
| | | Choreoathetosis, hypothyroidism, neonatal respiratory distress, NKX2-1 | - | 1 |
| | | Congenital heart defects, dysmorphic facial features, developmental delay, CDK13* | NEHI | 1 |

| | | | | |
|---|---|---|---|---|
| | | Growth retardation, impaired intellectual development, hypotonia, hepatopathy, IARS1* | - | 1 |
| | | Hemophagocytic lymphohistiocytosis, STXBP2* | - | 1 |
| | | Hyper IgE syndrome, IL6R | - | 1 |
| | | Hyper IgE syndrome, IL6ST | - | 1 |
| | | Intellectual developmental disorder, BRWD3* | - | 1 |
| | | Intellectual developmental disorder, DEPDC5* | - | 1 |
| | | Intellectual developmental disorder, SRRM2* | NEHI | 3 |
| | | Lissencephaly with complex brainstem malformation, MACF1* | - | 2 |
| | | Oculocerebrofacial syndrome (Kaufman type), UBE3B* | - | 1 |
| | | Pulmonary fibrosis and/or bone marrow failure, telomere-related, TERC | - | 1 |
| | | Pulmonary fibrosis and/or bone marrow failure, telomere-related, TERT | NEHI | 1 |
| 6 | Bronchiolitis obliterans | Chronic granulomatous disease, CYBA | - | 1 |
| | | Hyper IgE syndrome, DOCK8 | - | 1 |
| | | Immunodeficiency (CID), MCM4 | Obliterative bronchiolitis | 1 |
| | | Primary pulmonary hypertension, CAV1 | - | 1 |
| | | Respiratory infections, recurrent, and failure to thrive with or without diarrhoea, AGR2 | Chronic bronchiolitis | 2 |
| 7 | Suspected surfactant dysfunction disorder | Choreoathetosis, hypothyroidism, neonatal respiratory distress, NKX2-1 | - | 2 |
| 8 | Pulmonary alveolar Proteinosis | FARS1 associated disease, FARSA | PAP | 1 |
| | | Growth retardation, impaired intellectual development, hypotonia, hepatopathy, IARS1 | PAP | 1 |
| | | Immunodeficiency (CID), PNP | PAP | 1 |
| | | Interstitial lung and liver disease, MARS | PAP (3) | 37 |
| | | Lysinuric proteinuria, SLC7A7 | - | 1 |
| | | PAH with small patella syndrome, TBX4 | PAP | 1 |
| | | PAP due to immunodeficiency, OAS1 GoF | PAP | 2 |
| | | PAP due to phagocyte dysfunction, CSF2RA | PAP (9) | 19 |
| | | PAP due to phagocyte dysfunction, CSF2RB | - | 1 |
| 9 | Pulmonary hemorrhage | Autoinflammation and autoimmunity with immune dysregulation, COPA | Haemosiderosis | 4 |
| | | Familial cold inflammatory syndrome, NLRP3* | Haemosiderosis | 1 |
| | | Lacrimo-auriculo-dental-digital syndrome, FGF10 | - | 1 |
| | | PAH with small patella syndrome, TBX4 | Haemosiderosis (3) | 3 |
| 10 | Pulmonary hypertension | Alveolar capillary dysplasia with misalignment of pulmonary veins, FOXF1 | - | 4 |
| | | Brain small vessel disease, COL4A1 | - | 1 |
| | | Chr15q11 duplication syndrome* | - | 1 |
| | | Chr19q13 duplication syndrome* | - | 1 |
| | | Chr5q trisomy* | - | 1 |
| | | Chr9p monosomy* | - | 1 |
| | | Combined oxidative phosphorylation deficiency, SLC25A26 | - | 1 |

| | | | | |
|---|---|---|---|---|
| | | Congenital heart defects, dysmorphic facial features, developmental delay, CDK13* | - | 2 |
| | | Drayer syndrome, chr15q26 deletion | - | 1 |
| | | Facial dysmorphism, hypertrichosis, epilepsy, developmental delay, gingival overgrowth syndrome, KCNK4* | PH | 2 |
| | | Filamin A related ILD, FLNA | PH (3), Emphysema (2), alveolar hypoplasia | 10 |
| | | Jacobsen syndrome, chr11q23.3 deletion | - | 1 |
| | | Lymphatic malformation, PIEZO1 | PH | 1 |
| | | Multisystemic smooth muscle dysfunction syndrome, ACTA2 | - | 1 |
| | | Nonspherocytic hemolytic anemia, G6PD* | - | 1 |
| | | PAH with small patella syndrome, TBX4 | PH (3), alveolar hypoplasia (3) | 12 |
| | | Primary pulmonary hypertension, BMPR2 | PH, haemosiderosis | 2 |
| | | Primary pulmonary hypertension, COQ2 | - | 1 |
| | | Primary pulmonary hypertension, ENG1 | - | 2 |
| | | Primary pulmonary hypertension, KCNK3 | Haemosiderosis | 1 |
| | | Pulmonary hypertension/ pulmonary veno-occlusive disease, EIF2AK4 | - | 1 |
| | | STING-associated vasculopathy, infantile-onset, TMEM173 | - | 1 |
| | | Williams-Beuren syndrome, 7q11 deletion | PH | 1 |
| 11 | Interstitial pneumonia | Autoimmune disease, multisystemic, infantile-onset, STAT3 GoF | CPI, NSIP | 2 |
| | | Autoinflammation and autoimmunity with immune dysregulation, COPA | NSIP (3) | 3 |
| | | Choreoathetosis, hypothyroidism, neonatal respiratory distress, NKX2-1 | DIP, CP, NSIP | 3 |
| | | Chr9p monosomy* | DIP, NSIP | 2 |
| | | Chronic granulomatous disease, CYBA | DIP | 1 |
| | | Chronic granulomatous disease, NCF2 | DIP | 1 |
| | | Cystic lung disease with phagocyte dysfunction, CCR2 | DIP, CP | 2 |
| | | DiGeorge syndrome, chr22q11.2 deletion | NSIP (2) | 2 |
| | | Familial cold inflammatory syndrome, NLRP3* | NSIP (2) | 2 |
| | | Familial cold inflammatory syndrome, PLCG2 | DIP | 1 |
| | | Familial mediterranean fever, MEFV | DIP | 1 |
| | | FARS1 associated disease, FARSA | CP (3) | 3 |
| | | FARS1 associated disease, FARSB | CPI, CP (3) | 4 |
| | | Filamin A related ILD, FLNA | NSIP, UIP | 2 |
| | | FINCA syndrome, NHLRC2 | CP | 1 |
| | | Griscelli syndrome, RAB27A | NSIP | 1 |
| | | Immunodeficiency (CID), ATM | UIP | 1 |
| | | Immunodeficiency (CID), CD40* | UIP | 1 |
| | | Immunodeficiency (CID), HELLS* | CPI | 1 |
| | | Innate deficiency, ZNFX1 | CP, UIP | 2 |
| | | Interferonopathy, IFIH1 GoF* | CPI (2), DIP | 3 |
| | | PAH with small patella syndrome, TBX4 | NSIP | 1 |
| | | Peroxisome biogenesis disorder (Zellweger), PEX1 | DIP | 1 |
| | | Phagocyte deficiency, GATA2 | CP | 1 |

| | | | | |
|---|---|---|---|---|
| | | Pulmonary fibrosis and/or bone marrow failure, telomere-related, TERC | NSIP | 1 |
| | | Pulmonary fibrosis and/or bone marrow failure, telomere-related, TERT | UIP | 1 |
| | | STING-associated vasculopathy, infantile-onset, TMEM173 | NSIP (2) | 2 |
| | | Xp22.33 duplication syndrome* | CPI | 1 |
| 12 | Developmental arrest of the lung | Alveolar capillary dysplasia with misalignment of pulmonary veins, FOXF1 | ACD (16) | 16 |
| | | Lacrimo-auriculo-dental-digital syndrome, FGF10 | Acinar Dysplasia | 1 |
| | | Microphthalmia syndrome, STRA6 | ACD (2) | 2 |
| | | Xp22.33 duplication syndrome* | CAD | 1 |
| 13 | Lymphocyte/monocyte dominated ILD | Autoimmune disease, multisystemic, infantile-onset, STAT3 GoF | LIP (2) | 2 |
| | | Autoinflammation and autoimmunity with immune dysregulation, COPA | FB (7), LIP, DLH | 9 |
| | | Bronchiectasis with or without elevated sweat chloride, SCNN1A | FB | 1 |
| | | Choreoathetosis, hypothyroidism, neonatal respiratory distress, NKX2-1 | FB | 1 |
| | | Chronic granulomatous disease, CYBA | GLILD | 1 |
| | | Chronic granulomatous disease, CYBB | GLILD | 1 |
| | | DiGeorge syndrome, chr22q11.2 deletion | FB | 1 |
| | | Immunodeficiency (CID), IL2RG | FB | 1 |
| | | Immunodeficiency (CVID), IRF2BP2* | FB | 1 |
| | | Immunodeficiency (CVID), LRBA | FB | 1 |
| | | Immunodeficiency (CVID), TNFRSF13B | GLILD | 1 |
| | | Pulmonary fibrosis and/or bone marrow failure, telomere-related, TERT | FB | 1 |
| | | STING-associated vasculopathy, infantile-onset, TMEM173 | FB | 1 |
| 14 | Pulmonary interstitial glycogenosis | Microphthalmia syndrome, RARB | PIG | 1 |
| 15 | Abnormal lung vessels | Diets-Jongmans syndrome, KDM3B | PCH | 1 |
| | | Primary pulmonary hypertension, CAPNS1 | PCH | 1 |
| | | Pulmonary hypertension/ pulmonary veno-occlusive disease, EIF2AK4 | PCH | 1 |
| | | Xp22.33 duplication syndrome* | PCH | 1 |
| 16 | Undefined ILD | Autoimmune disease, multisystemic, infantile-onset, STAT3 GoF | Alveolar hypoplasia | 2 |
| | | Carnitine-acylcarnitine translocase deficiency, SLC25A20 | - | 1 |
| | | Choreoathetosis, hypothyroidism, neonatal respiratory distress, NKX2-1 | - | 12 |
| | | Chr5q trisomy* | - | 1 |
| | | Chronic granulomatous disease, CYBB | - | 2 |
| | | Developmental and epileptic encephalopathy, PCDH19* | Fibrosis | 1 |
| | | Familial cold inflammatory syndrome, PLCG2 | Alveolar hypoplasia | 1 |
| | | Familial erythrocytosis/ Rothshild, HBB | Alveolar hypoplasia | 1 |
| | | FARS1 associated disease, FARSB | Alveolar hypoplasia | 2 |
| | | Filamin A related ILD, FLNA | Alveolar hypoplasia | 3 |
| | | FINCA syndrome, NHLRC2 | - | 1 |
| | | Hemoglobinopathy, HBA2 | Alveolar hypoplasia | 1 |
| | | Hyper IgE syndrome, DOCK8 | - | 1 |
| | | Immunodeficiency (CID), ADA | Fibrosis | 1 |
| | | Immunodeficiency (CID), ATM | - | 1 |
| | | Immunodeficiency (CID), ATR* | - | 1 |

| | | | | |
|---|---|---|---|---|
| | | Immunodeficiency (CID), IL2RG | - | 1 |
| | | Immunodysregulation, polyendocrinopathy and enteropathy, FOXP3 | - | 1 |
| | | Innate deficiency, ZNFX1 | - | 2 |
| | | Interferonopathy, IFIH1 GoF* | Alveolar hypoplasia | 1 |
| | | Kabuki syndrome, KDM6A* | Emphysema | 1 |
| | | Lymphangioleiomyomatosis, TSC1 | Fibrosis | 1 |
| | | Multisystemic smooth muscle dysfunction syndrome, ACTA2 | - | 1 |
| | | PAH with small patella syndrome, TBX4 | Fibrosis | 1 |
| | | PAP due to phagocyte dysfunction, CSF2RA | - | 1 |
| | | Pulmonary fibrosis and/or bone marrow failure, telomere-related, DKC1 | - | 1 |
| | | Pulmonary fibrosis and/or bone marrow failure, telomere-related, RTEL1 | - | 1 |
| | | Pulmonary fibrosis and/or bone marrow failure, telomere-related, TERT | - | 1 |
| | | STING-associated vasculopathy, infantile-onset, TMEM173 | Alveolar hypoplasia, eosinophilic pneumonitis, fibrosis, chronic bronchiolitis | 12 |
| | | Wolfram syndrome, WFS1* | - | 1 |
| | | Xp22.33 duplication syndrome* | - | 1 |
| | **ILD with genetic diagnosis, affecting the lungs only** | | | **170** |
| 4 | Alveolar microlithiasis (PAM) | Alveolar microlithiasis | PAM (3) | 3 |
| 7 | Suspected surfactant dysfunction disorder | Surfactant dysfunction disorder, ABCA3 | Non-dx, eosinophilia | 52 |
| | | Surfactant dysfunction disorder, SFTPA1 | - | 1 |
| | | Surfactant dysfunction disorder, SFTPB | Non-dx (2) | 17 |
| | | Surfactant dysfunction disorder, SFTPC | Fibrosis, emphysema, chronic bronchiolitis | 27 |
| 8 | Pulmonary alveolar Proteinosis | Surfactant dysfunction disorder, ABCA3 | PAP (2) | 2 |
| | | Surfactant dysfunction disorder, SFTPB | PAP | 1 |
| | | Surfactant dysfunction disorder, SFTPC | PAP (6) | 8 |
| 11 | Interstitial Pneumonia | Surfactant dysfunction disorder, ABCA3 | CPI (30), DIP (3), NSIP (5), UIP | 39 |
| | | Surfactant dysfunction disorder, SFTPB | CPI (3), DAD, NSIP | 5 |
| | | Surfactant dysfunction disorder, SFTPC | CPI (11), CP, NSIP (2) | 14 |
| 13 | Lymphocyte/monocyte dominated ILD | Surfactant dysfunction disorder, SFTPC | LIP | 1 |
| | **ILD without genetic diagnosis** | | | **426** |
| 2 | Pulmonary hypoplasia | Pulmonary hypoplasia | Pulmonary hypoplasia | 3 |
| 3 | Persistent tachypnea of infancy (PTI/NEHI)) | Persistent tachypnea of infancy (PTI/NEHI) | NEHI (17), non-dx (5), chronic bronchiolitis (7), alveolar hypoplasia (2) | 137 |
| 6 | Bronchiolitis obliterans | Bronchiolitis obliterans (BO) without suspected cause | Constrictive bronchiolitis | 5 |
| 7 | Suspected surfactant dysfunction disorder | Undefined ILD in almost (30-36 weeks) mature; no/low SP-C biochem | Alveolar hypoplasia, non-dx | 9 |
| | | Undefined ILD in mature neonate; no/low SP-C biochem | Alveolar hypoplasia, chronic bronchiolitis, non-dx | 31 |
| | | Undefined ILD in NON neonate; no/low SP-C biochem | - | 4 |
| 8 | Pulmonary alveolar Proteinosis (PAP) | Pulmonary alveolar proteinosis | PAP (5) | 11 |
| 9 | Pulmonary hemorrhage | Idiopathic pulmonary haemosiderosis (IPH) | DAH, Haemosiderosis (5) | 10 |

| | | | | |
|---|---|---|---|---|
| | | Pulmonary hemorrhage | - | 6 |
| 10 | Pulmonary hypertension | Pulmonary hypertension | PH (5), alveolar hypoplasia, non-dx | 39 |
| 11 | Interstitial pneumonia | Chronic pneumonitis of infancy | CPI (19) | 19 |
| | | Desquamative interstitial pneumonia | DIP (6) | 6 |
| | | Diffuse alveolar damage (acute interstitial pneumonia) | DAD (4) | 4 |
| | | Lipoid pneumonitis, Cholesterol pneumonia | CP (2) | 2 |
| | | Nonspecific interstitial pneumonia | NSIP (18), mixed pattern NSIP/DIP (6), NSIP/PAP (3) | 27 |
| | | Usual interstitial pneumonitis | UIP | 1 |
| 12 | Developmental arrest of the lung | Alveolar capillary dysplasia | ACD (5) | 5 |
| | | Alveolar capillary dysplasia, misalignment of pulmonary veins | ACD (6) | 6 |
| | | Congenital alveolar dysplasia | CAD (7) | 7 |
| 13 | Lymphocyte/monocyte dominated ILD | Diffuse lymphoid hyperplasia | DLH | 1 |
| | | Follicular bronchitis/bronchiolitis | FB (7) | 7 |
| | | Granulomatous lung disease | GLILD | 1 |
| | | Lymphocytic interstitial pneumonia | LIP (5) | 5 |
| 14 | Pulmonary interstitial glycogenosis (PIG) | Pulmonary interstitial glycogenosis (PIG) | PIG (14) | 14 |
| 15 | Abnormal lung vessels | Pulmonary capillary hemangiomatosis | PCH (14) | 14 |
| | | Veno-occlusive disease | PVOD | 1 |
| 16 | Undefined ILD | Alveolar simplification/hypoplasia | Alveolar hypoplasia (2) | 2 |
| | | Cystic lung disease, unknown etiology | - | 1 |
| | | Emphysema | Emphysema (3) | 4 |
| | | Eosinophilic pneumonitis | Eosinophilic pneumonitis | 1 |
| | | Fibrotic lung disease, unknown etiology | Fibrosis (3) | 7 |
| | | Undefined ILD in almost (30-36 weeks) mature | - | 6 |
| | | Undefined ILD in mature neonate | Non-dx | 17 |
| | | Undefined ILD in NON neonate | Chronic Bronchiolitis (2) | 13 |

**Tab. S4: Occasional genetic testing in ILD with identified cause/ lung injury or a convincing diagnosis of systemic disease.** Genetic analysis was initiated on request of the attending physician, even if ILD etiology was identified with sufficient clinical certainty. Genetic identifier lists the affected gene with their occurrence, if identified more than once.

| | Clinical entities | N = | Genetically analyzed, N = | To exclude ILD, affecting the lung only | To confirm systemic disease | Incidental findings | Genetic identifier |
|---|---|---|---|---|---|---|---|
| | **ILD with identified cause/lung injury** | **419** | **153 (36.5%)** | **145 (94.8%)** | **-** | **8 (5.2%)** | |
| 1 | Abnormal lung development in pre-term | 176 | 89 (50.6%) | 87 | - | 2 | 6q23 del, distal trisomy 16q (identified later in 2 infants with BPD) |
| 4 | Pulmonary hypoplasia, compression/ displacement | 35 | 15 (42.9%) | 14 | - | 1 | *FKBP10* *(identified in an infant with pulmonary hypoplasia due to chylothorax* |
| 7 | Exposure-related ILD | 130 | 28 (21.5%) | 24 | - | 4 | 22q11.2 del, 2q15del, Monosomy 21, 5q31.3 del (identified later in 4 ILD from chronic aspirations) |
| 9 | Bronchiolitis obliterans, post-infectious (PIBO) | 78 | 21 (26.9%) | 20 | - | 1 | *IFIH1* Loss of Function (identified in a patient with PIBO) |
| | **ILD with convincing diagnosis of systemic disease** | **365** | **197 (54.0%)** | **39 (19.8%)** | **158 (81.2%)** | - | |
| 17 | Syndromic disorders (86), cardiovascular phenotype (82), autoantibody positivity (42), suspected inborn error of immunity (31), langerhans cell histiocytosis/ sarcoidosis (26), lymphatic vessel involvement (18), neurological phenotype (16), autoimmune PAP (15), hermansky pudlak (10), Niemann Pick (9), Noonan syndrome (8), lysinuric proteinuria, musculoskeletal phenotype (5), others (11) | 365 | 197 | 39 | 158 | - | 10p13 del, 11q23 del, 21 Trisomy (59), 22 Trisomy (4), 22q11.2 del (5), 3q26 dup, 4p16.3 del, *ABCC9* (2), *ABCD1* (2), *ACVRL1, ADA, AP3B1* (7), *BMPR2, CD40, CDK13, CHD7, COL3A1, COPA, CREBBP, CTLA4, DKC1 (2), ENG1 (2), FAT4, FBN1, FLCN, FOXP3* (2), *GATA2, GJC2, GNPTAB, HBB, HPS1* (2), *IDS, IFIH1, IKBKG, ITGA-3* (2), *KAT6B, MAPK1, MECP2* (2), *MEFV, NAGLU* (2), *NCF4, NHEJ1, NHLRC2, NKX2-1, NPC1, NPC2* (3), *PBX1, PIEZO1, POLR3A* (2), *PTPN11* (2), *RASA1, RIT1, SGCB, SGSH, SH3TC2, SLC7A7* (3), *SMAD4* (2), *SMPD-1, STAT4, STXPB2, TERT, TNFRSF13B, TSC1, TTC37, UBR1, WAS,* X duplication (3), *ZNFX1 (2)* |

**Tab. S5: Most frequently observed non-pulmonary organ manifestation in patients with ILD (N = 1,686).** The non-pulmonary organ manifestation was described by standardized human phenotype ontology (HPO) terms, which were linked to major organ groups.

| **Major organ groups** | **Top five used HPO terms per organ (N = )** |
|---|---|
| Autoimmune features | HP:0003493: Antinuclear antibody positivity(16); HP:0020050: Anti-granulocyte-macrophage colony stimulating factor antibody positivity (15); HP:0003453: Antineutrophil antibody positivity (9); HP:0025343: Lupus anticoagulant (8); HP:0003613: Antiphospholipid antibody positivity (6) |
| Lymphatic features | HP:0001744: Splenomegaly (34); HP:0002955: Granulomatosis (20); HP:0002716: Lymphadenopathy (20); HP:0010310: Chylothorax (13); HP:0001789: Hydrops fetalis (8); ); |
| Immunodeficiency features | HP:0004313: Decreased circulating antibody level (38); HP:0100806: Sepsis (27); HP:0004432: Agammaglobulinemia (11); HP:0005681: Juvenile rheumatoid arthritis (11); HP:0001974: Leukocytosis (11); |
| Vascular features | HP:0004890: Elevated pulmonary artery pressure (288); HP:0011726: Persistent fetal circulation (38); HP:0002239: Gastrointestinal hemorrhage (9); HP:0001028: Hemangioma (9); HP:0000421: Epistaxis (8); |
| Endocrine features | HP:0000821: Hypothyroidism (71); HP:0000836: Hyperthyroidism (10); HP:0000872: Hashimoto thyroiditis (8); HP:0000846: Adrenal insufficiency (7); HP:0000829: Hypoparathyroidism (5) |
| Syndromic features (incl. head and neck) | HP:0000252: Microcephaly (36); HP:0000365: Hearing impairment (31); HP:0500049: Retinopathy of prematurity (25); HP:0001999: Abnormal facial shape (21); HP:0000407: Sensorineural hearing impairment (11); |
| Dermal features (incl. hair and nails) | HP:0001217: Clubbing (50); HP:0001047: Atopic dermatitis (25); HP:0000961: Cyanosis (17); HP:0000964: Eczematoid dermatitis (13); HP:4000054: Exanthem (12); |
| Muscular features | HP:0001252: Hypotonia (117); HP:0000776: Congenital diaphragmatic hernia (18); HP:0000023: Inguinal hernia (17); HP:0012378: Fatigue (9); HP:0001537: Umbilical hernia (9); |
| Cardiac features | HP:0001631: Atrial septal defect (125); HP:0001643: Patent ductus arteriosus (111); HP:0001655: Patent foramen ovale (89); HP:0001629: Ventricular septal defect (48); HP:0005180: Tricuspid regurgitation (37) |
| Gastroenterological/ pancreatic features | HP:0002020: Gastroesophageal reflux (85); HP:0002013: Vomiting (36); HP:0002014: Diarrhea (30); HP:0002027: Abdominal pain (24); HP:0002608: Celiac disease (19) |
| Hepatic (incl. gall bladder) features | HP:0002240: Hepatomegaly (59); HP:0002904: Hyperbilirubinemia (20); HP:0001410: Decreased liver function (16); HP:0001396: Cholestasis (14); HP:0002910: Elevated circulating hepatic transaminase concentration (14) |
| Hematological features | HP:0001903: Anemia (87); HP:0001873: Thrombocytopenia (51); HP:0001875: Neutropenia (19); HP:0001891: Iron deficiency anemia (18); HP:0001880: Eosinophilia (12); |
| Neurological features | HP:0001263: Global developmental delay (87); HP:0001250: Seizure (58); HP:0001270: Motor delay (37); HP:0002361: Psychomotor deterioration (30); HP:0012758: Neurodevelopmental delay (29); |
| Nephrotic/ urogenital features | HP:0000083: Renal insufficiency (23); HP:0000121: Nephrocalcinosis (16); HP:0000093: Proteinuria (14); HP:0000126: Hydronephrosis (13); HP:0012622: Chronic kidney disease (11); |
| Skeletal features | HP:0002829: Arthralgia (42); HP:0000767: pectus excavatum (42); HP:0002650: Scoliosis (26); HP:0001385: Hip dysplasia (9); HP:0001382: Joint hypermobility (9); |

**Fig. S1: Results of genetic investigation in children with ILD, unknown aetiology, N = 902.** In 476 (52.8%) patients, a genetic etiology was identified. Then patients were re-categorized into three aetiological cohorts: ILD with genetic diagnosis (disease-causing variants in *SLC34A2, ABCA3, SFTPA, SFTPB*, and *SFTPC*), ILD with genetic diagnosis of systemic disease (details of final diagnoses, Tab. 2, Tab. 1S), and ILD without genetic diagnosis. Frequency of genetic etiology was listed if identified more than once, genes identified only once were summarized under "Other" and are listed in Tab. 2 with their final diagnosis in Tab. 1S.

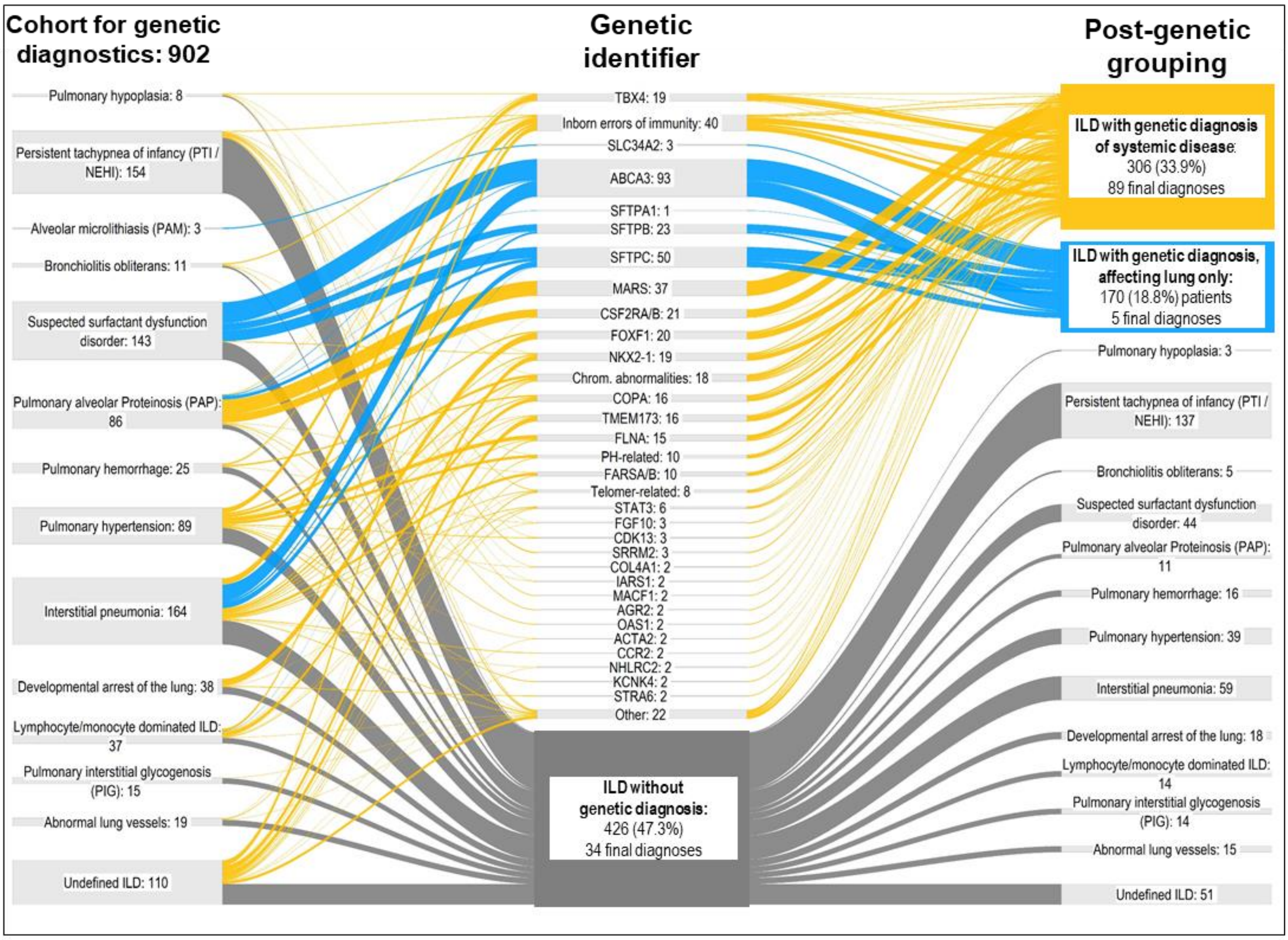
